# From Evidence to Data Framework: Decision Factors and Structured Data for AI-Driven Clinical Decision Support Systems in Offloading Footwear

**DOI:** 10.1101/2025.08.20.25334118

**Authors:** Kunal Kumar, Muhammad Ashad Kabir, Luke Donnan, Sayed Ahmed, Martin Nguyen

**Affiliations:** School of Computing, Mathematics and Engineering, Charles Sturt University, Bathurst, Australia; School of Allied Health, Exercise and Sports Sciences, Charles Sturt University, Albury, Australia; Foot Balance Technology Pty Ltd, Castle Hill, Australia

**Keywords:** Diabetic foot ulcer, clinical decision support systems, offloading footwear, orthotics, ulcer healing, plantar pressure, footwear adherence

## Abstract

**Background:** Diabetes-related foot ulcers (DFUs) are a serious complication of diabetes, often resulting in infection, amputation, or even mortality. Offloading footwear is a key intervention that promotes ulcer prevention and healing by reducing pressure on affected areas of the foot. However, patient adherence to prescribed footwear remains low. Current clinical guidelines, such as those from the International Working Group on the Diabetic Foot (IWGDF), offer general recommendations but lack a standardized and structured framework. Furthermore, they do not support personalized footwear prescription or integration into clinical decision support systems (CDSSs) and artificial intelligence (AI) applications.

**Objective:** The primary objective of this study was to identify and systematically categorise the key factors influencing footwear prescription and design for patients with DFUs. A secondary objective was to integrate these factors into a structured framework aligned with HL7 Fast Healthcare Interoperability Resources (FHIR) standards, with the goal of informing the development of AI-powered CDSSs.

**Methods:** A systematic scoping review was conducted in accordance with the PRISMA-ScR guidelines. Seventeen academic databases and Google Scholar were searched for relevant studies published between 2014 and 2025. Eligible studies included original research involving adults with diabetes or diabetic foot ulcers (DFUs) and focused on offloading footwear or orthotic interventions. Data were extracted and categorized using the WHO Dimensions of Adherence to Long-Term Therapies framework and mapped to FHIR resources using standardized terminologies such as SNOMED CT, and LOINC. Where existing standards were insufficient, custom FHIR extensions were proposed.

**Results:** A total of 81 studies met the inclusion criteria, encompassing data from 5,001 participants. Key outcome measures across the studies included plantar pressure reduction (n = 55), adherence (n = 27), gait and balance (n = 20), and ulcer recurrence (n = 13). The review identified 90 unique decision factors influencing footwear prescription, classified according to the five WHO dimensions of adherence to long-term therapies: patient-related, condition-related, therapy-related, socioeconomic, and healthcare system– related factors. Of the 90 factors identified, 48 were categorized as input variables and 42 as output variables. The review found that only 16/90 of the decision factors could be mapped directly to existing FHIR standards, while the remaining 74/90 required custom extensions—particularly those related to detailed footwear attributes and socioeconomic patient data. This is because of the unique nature of the domain as the result of lack of standardized definition related to the domain.

**Conclusions:** This study introduces the first structured and interoperable framework for prescribing offloading footwear in the management of diabetic foot ulcers (DFUs). By aligning key decision factors with HL7 FHIR standards and the WHO Dimensions of Adherence, the framework enables seamless integration into electronic health records and supports the development of AI-driven clinical decision support systems (CDSSs). Future work should prioritize expert validation of the identified factors, implementation studies, and real-world testing to evaluate the framework’s usability, clinical relevance, and impact on patient outcomes.

## Introduction

Diabetes-related foot ulcers (DFUs) are a serious complication of diabetes, affecting approximately 15% of diabetes patients in their lifetime [1]. While repetitive mechanical stress on the foot is the primary cause of DFUs, other conditions such as diabetic peripheral neuropathy (DPN) and peripheral arterial disease (PAD) further impair healing and increase the risk of infections [2]. If left untreated, DFUs can lead to severe complications, including amputations, and higher mortality rates[3]. Despite developments in DFU care, significant gaps persist in optimizing clinical interventions such as offloading footwear [4, 5].

Appropriate offloading footwear is a key intervention for reducing excessive plantar pressure (PP) and redistributing mechanical stress, subsequently preventing DFU formation and allowing ulcers to heal [6]. The efficacy of offloading footwear is well-documented, with studies demonstrating significant reductions in PP and slower DFU progression [7–9]. Additionally, research evidence suggests that around 75% of DFUs can be prevented with offloading footwear [10]. Despite these benefits, the effectiveness of footwear is impaired due to lack of adherence by patients. As a result, approximately 40% of DFU patients with low adherence experience a recurrence within one year of healing, and 65% within five years [11]. Various studies have found different reasons behind the lack of adherence. For instance, some studies have indicated that patients often underutilize offloading footwear due to aesthetic concerns, discomfort, or a lack of perceived benefits [12–14]. Another study found that patients often struggle to integrate prescribed footwear into their daily routines [15]. Furthermore, factors such as fit, lifestyle constraints, and cultural beliefs are also been shown to influence adherence [16]. While specific findings behind low adherence vary among studies, non-clinical factors play a major role in this. As a result, research suggests a patient-centric approach that accounts for both pathological and non-pathological factors to improve adherence [4].

Current guidelines, including those from the International Working Group on the Diabetic Foot (IWGDF) [17], and Australian guidelines [18] provide evidence-based recommendations for DFU prevention. While these guidelines outline key features of DFU offloading footwear, such as appropriate fit, cushioning, and rigidity to offload high-pressure regions of the foot, they do not account for specific non-clinical factors. A systematic review of DFU guidelines has highlighted a lack of standardized and structured frameworks for footwear prescription [19]. Current approaches often emphasizing general recommendations rather than defining a comprehensive set of attributes necessary for effective prescription [19]. Footwear prescription for DFU patients is a complex process involving various clinical and non-clinical factors, as well as personalization based on patient needs. In addition to clinical factors, effective footwear prescription requires consideration of non-clinical factors such as patient expectations, living environments, and footwear aesthetics to enhance adherence [4]. This added complexity underscores the need for standardization to improve clinical efficiency and ensure optimal patient outcomes.

Despite the well-established role of footwear in DFU prevention, a significant gap exists in the literature regarding the standardization of factors needed for making footwear decisions. Existing studies primarily focus on specific footwear components, such as insole materials or rocker profiles, but lack a comprehensive framework defining the essential decision factors for prescription. This absence of standardization leads to variability in footwear recommendations and affects the development of optimal prescriptions for individual patients. This scoping review aims to address this gap by systematically identifying key decision factors and developing a standardized framework to guide data collection in practice. By synthesizing existing literature, this study will establish a foundation for improved data collection and the development of artificial intelligence (AI)-driven clinical decision support systems (CDSS) for footwear prescription. These CDSSs can assist healthcare providers in making evidence-based decisions and potentially improve patient outcomes such as reducing ulcer incidence, promoting efficient healing, and improving adherence.

### Study Objectives

- To identify and categorize key factors used in DFU footwear prescription decisions across clinical studies.
- To establish a structured framework of data elements for DFU footwear prescription, providing a foundation for AI-driven clinical decision support systems.

## Methods

### Study Design

This study conducts a systematic scoping review to explore quantitative and qualitative studies that identify factors influencing the prescription of offloading footwear for diabetic foot ulcers (DFUs), and to summarize these factors along with appropriate data types and sample values. A scoping review is the most feasible study design to address the broad objectives of the study and map diverse empirical evidence. This review was guided by the methodologies of Arksey and O’Malley [20], Munn et al [21], and Pollock et al [22], and follows the Preferred Reporting Items for Systematic Review and Meta-analysis extension for Scoping Reviews (PRISMA-ScR) guidance [23]. The study does not assess methodical quality of the included studies because it does not align with the research objectives. The review was registered prospectively on the open science framework for systematic reviews [24].

### Search Strategy

Search was facilitated through Primo Library Search tool. Primo contains the following list of databases: DOAJ Directory of Open Access Journals, EBSCOhost, ScienceDirect, Food Science Source, IngentaConnect, Ovid, Open Access Digital Library, ProQuest, PubMed, ROAD: Directory of Open Access Scholarly Resources, SAGE, Web of Science, SciTech, Scopus, Single Journals, Springer, and Wiley. Additionally, Google Scholar was searched to identify studies not included in Primo. The initial search strategy consisted of broad terms synonymous with “foot ulcer”, “orthotics‟, “adherence” and “offloading footwear”. Further analysis of keywords from extracted articles was used to develop the final search strategy, as displayed in Table 1.

**Table 1:** Search keywords.

| Field | Search Terms | Boolean |
| --- | --- | --- |
| 1 | "diabetes" OR "diabetic" OR "diabetic foot ulcer*" OR "diabetes-related foot ulcer" OR "foot ulcer" OR "ulcer", OR "adherence", OR "compliance" | AND |
| 2 | "ulcer prevention" OR "offloading", OR "offload*" OR "plantar pressure*" OR "treatment" OR "therapy" | AND |
| 3 | "footwear" OR , "shoe*" OR "insole" OR "orthotics" OR "ortho*" OR "pedorthic" |  |

### Eligibility Criteria

The reviews eligibility criteria is guided by the PICO framework [25], which is commonly used to guide research questions in the medical field. Studies were selected based on the following inclusion criteria: (1) population: adults aged 18 years and over, and a diagnosis of diabetes or DFU; (2) intervention: conventional offloading footwear or orthotics; (3) comparator: another treatment option or no treatment or no comparator; (4) outcome: Ulcer prevention (plantar pressure reductions, gait or balance, temperature changes, ulcer incidences, ulcer recurrence, or patient adherence, or other outcomes identified during data extraction); (5) type of study: the review included both epidemiological and experimental study designs including randomized controlled trials, non-randomized control trials, before and after studies, quasi-experimental, cross-sectional studies, prospective and retrospective studies, case study, and observational studies. Additionally, the study placed specific focus on recent research articles that were published between 2014 and 2025.

Excluded studies included those that were: (1) not accessible in full text, (2) review articles, (3) not original research including case series, case reports, and expert commentaries, (4) grey literature and thesis databases (5) did not focus on DFU population in isolation, (6) did not focus on offloading footwear in isolation, (7) did not focus on offloading footwear and orthotics in isolation, or (8) focused on removable cast walker(RCWs), (9) focused on specialized, smart, or intelligent footwear and insoles, or (10) did not sufficiently discuss the study outcomes.

### Data Extraction and Charting

Study screening and data extraction was conducted by a team of researchers (KK, AK, LD, MN, SA). Potential studies were imported to Covidence for screening where two reviewers (KK and MN) independently screened each articles title and abstract. Articles marked for inclusion by both reviewers were moved to full-text screening, which was subsequently completed by the two reviewers (KK and MN). Any conflicts during the screening process were resolved by discussion among the research team. After screening, one reviewer (KK) extracted study characteristics including purpose of the study, year of publication, country of publication, study design, and population demographics using NVivo software. Additionally, the reviewer conducted a framework based thematic analysis with NVivo software. The thematic was used the WHO Dimensions of Adherence to Long-Term Therapies, systematically identifying and categorizing all data elements of footwear prescription into patient-related, condition-related, socioeconomic, therapy-related, and health-system-related factors. Subcategories and associated values will be thematically coded in NVivo under one of these categories. Furthermore, the study performed concept alignment with Fast Healthcare Interoperability Resources (FHIR) resources, where each data element was mapped to concepts in standardized terminology systems such as SNOMED, ICD-10, UCUM, and LOINC. Custom extensions were created for data elements that do not match existing concepts. The data extracted by KK was independently reviewed by two team members (LD, SA), where conflicts and disagreements will be resolved by a third reviewer (AK).

### Data Extraction Frameworks

Data extraction was guided by two frameworks, “WHO Dimensions of Adherence to Long-Term Therapies” and “Fast Healthcare Interoperability Resources (FHIR)”. The WHO Dimensions guided the thematic analysis while the FHIR resources was used for mapping the data elements to FHIR code systems.

#### WHO Dimensions of Adherence to Long-Term Therapies

The findings of the 2003 WHO report “Dimensions of Adherence to Long-Term therapies” was used to structure each studies data [26]. The report categorizes data elements into five key categories, namely, patient-related, condition-related, socioeconomic, therapy-related, and health-system related. The WHO report did not provide explicit definitions for the five domains, rather it lists factors that it believed to fall in that category.

#### Fast Healthcare Interoperability Resources (FHIR)

FHIR is the latest standard developed by Health Level 7 (HL7) to enhance information exchange among healthcare systems. These standards are essential for the adoption of electronic health record systems (EHRs) and their integration with CDSSs. FHIR resources can be mapped to real-world data elements, enabling structured and standardized data representation. This study categorized each identified data element into appropriate FHIR categories by mapping them to FHIR terminologies. The mappings were determined using standardized terminology databases supported by FHIR, such as SNOMED CT, RxNorm, and LOINC.

### Ethical Considerations

Since this research was solely based on literature and did not engage any research participants or subjects, no formal ethics approval was required from the Charles Sturt University Human Research Ethics Committee.

## Results

### Study Selection

A total of 1,611 studies were identified across various academic journals. After removing duplicates using the Covidence tool, 929 unique records remained eligible for screening. Following the title and abstract screening, 125 articles were deemed eligible for full-text review. An additional 40 papers were identified at this stage through the snowballing technique, resulting in 165 articles eligible for full-text screening. After full-text assessment, 84 studies were excluded for reasons such as inaccurate outcomes, population mismatch, and inappropriate interventions. The PRISMA flowchart is shown in Figure 1.

**Figure 1:**
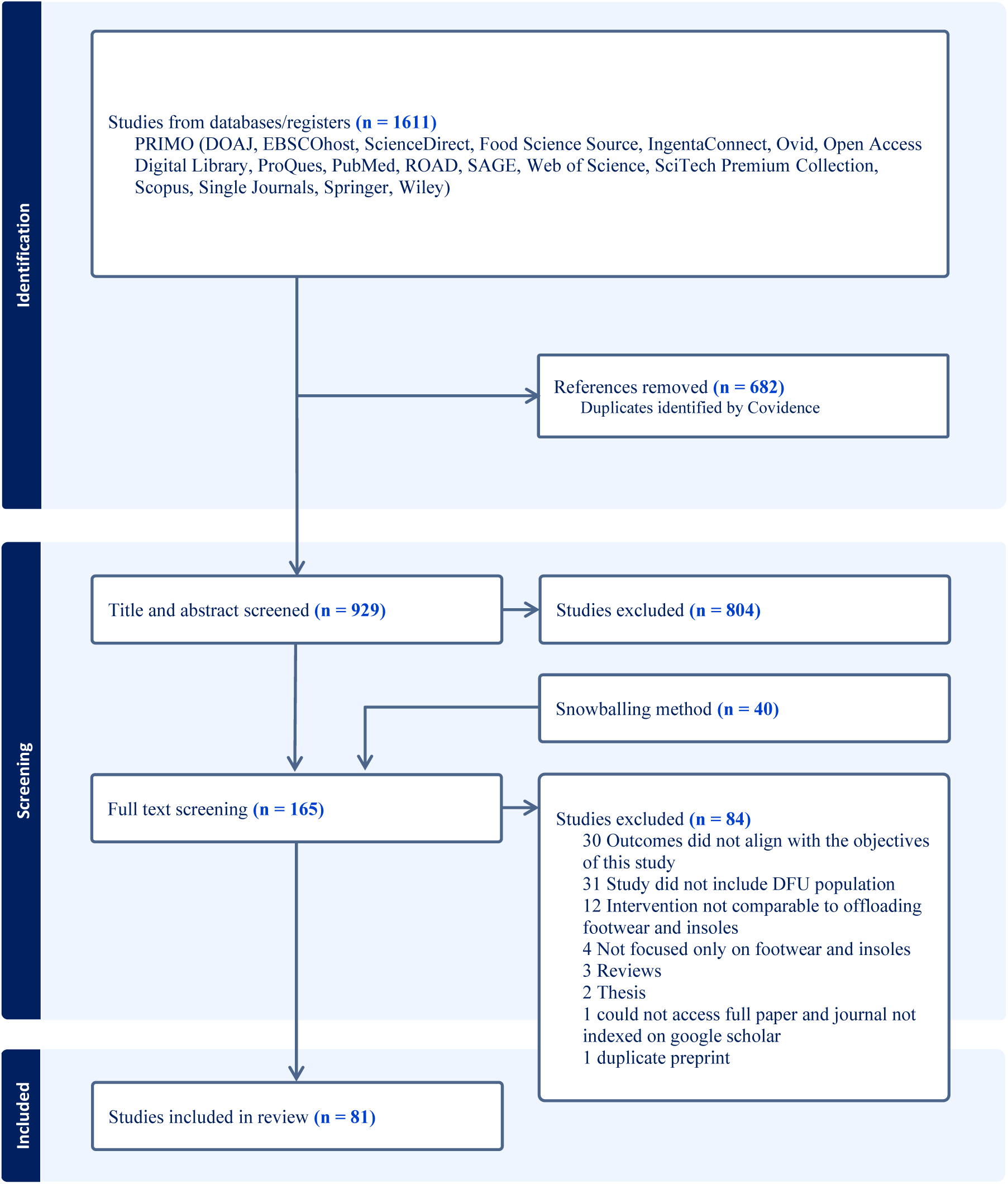
PRISMA (Preferred Reporting Items for Systematic Reviews and Meta-Analyses) diagram.

### Study Characteristics

A total of 5,001 participants were included across the studies. The mean age was 46.01 years (SD 10.56), ranging from 18 to 84 years, indicating a predominance of middle-aged adults. Gender was inconsistently reported, with 1,813 participants (36.2%) identified as male and 1,007 (20.1%) as female. Diabetes type was not consistently described; among those with available data, 298 participants (6.0%) had type 1 diabetes and 1,729 (34.6%) had type 2 diabetes. Glycemic control was reported in a subset of studies, with a mean value of 56.91 mmol/mol (SD 14.93). Mean BMI was 30.0 kg/m² (SD 5.33), suggesting most participants were in the obese range. Mean weight was 80.76 kg (SD 15.37), and mean height was 166.38 cm (SD 8.46). This demographics information is presented in Table 2. Demographic variables such as gender and diabetes classification were frequently underreported, limiting subgroup analyses and the ability to assess representativeness across study populations.

**Table 2:** Participant Demographics.

| Category | Participants (N = 5001) |
| --- | --- |
| Age | 46.01 ± 10.56 |
| Age Range | 18 - 84 |
| **Gender:** |  |
| *Male* | 1813 |
| *Female* | 1007 |
| **Diabetes:** |  |
| *Type 1* | 298 |
| *Type 2* | 1729 |
| *HbA1c (mmol/mol)* | 56.91 ± 14.93 |
| **BMI** | 30 ± 5.33 |
| **Weight (kg)** | 80.76 ± 15.37 |
| **Height (cm)** | 166.38 ± 8.46 |

Assessment of study origin by country indicated that the Netherlands is identified as the leading contributor, with 20 studies conducted in this setting [15, 27–45]. The United Kingdom [12, 46–53] and Australia [13, 54–60], each contributed 8 studies respectively. The United States contributed with 7 studies [12, 53, 61–65], while Spain has 5 studies [66–70]. Iran [52, 65, 71, 72], China [73–76], and Thailand [77–80] contributed 4 studies each. Germany [38, 81, 82], Malta [83–85], and Sweden [14, 86, 87] each contributed 3 studies. Several countries had 2 studies each, including India [88, 89], Italy [90, 91], Portugal [92, 93], and Egypt [94, 95]. The following countries contributed one study each: Jordan [96], Canada [97], Indonesia [98], Colombia [99], Singapore [100], Israel [101], and Fiji [102].

Four studies were conducted across multiple countries. One study was conducted between the Netherlands and Germany [38], another study was jointly conducted by the United States and the United Kingdom [12], two studies were conducted in collaboration between Iran and the United Kingdom [52, 65], and one study was conducted in United Kingdom and Germany [82]. These overlaps highlight a small but important trend toward international collaboration in technology-enabled diabetes and mental health care research.

Figure 2 presents the geographic distribution of study origins, revealing a clear imbalance between developed and developing countries. Western nations such as the Netherlands, the United Kingdom, and Australia dominate research contributions. In contrast, only a few developing countries are represented, and their contributions remain limited. This underscores the need for greater support and inclusion of developing nations in research within this field.

**Figure 2:**
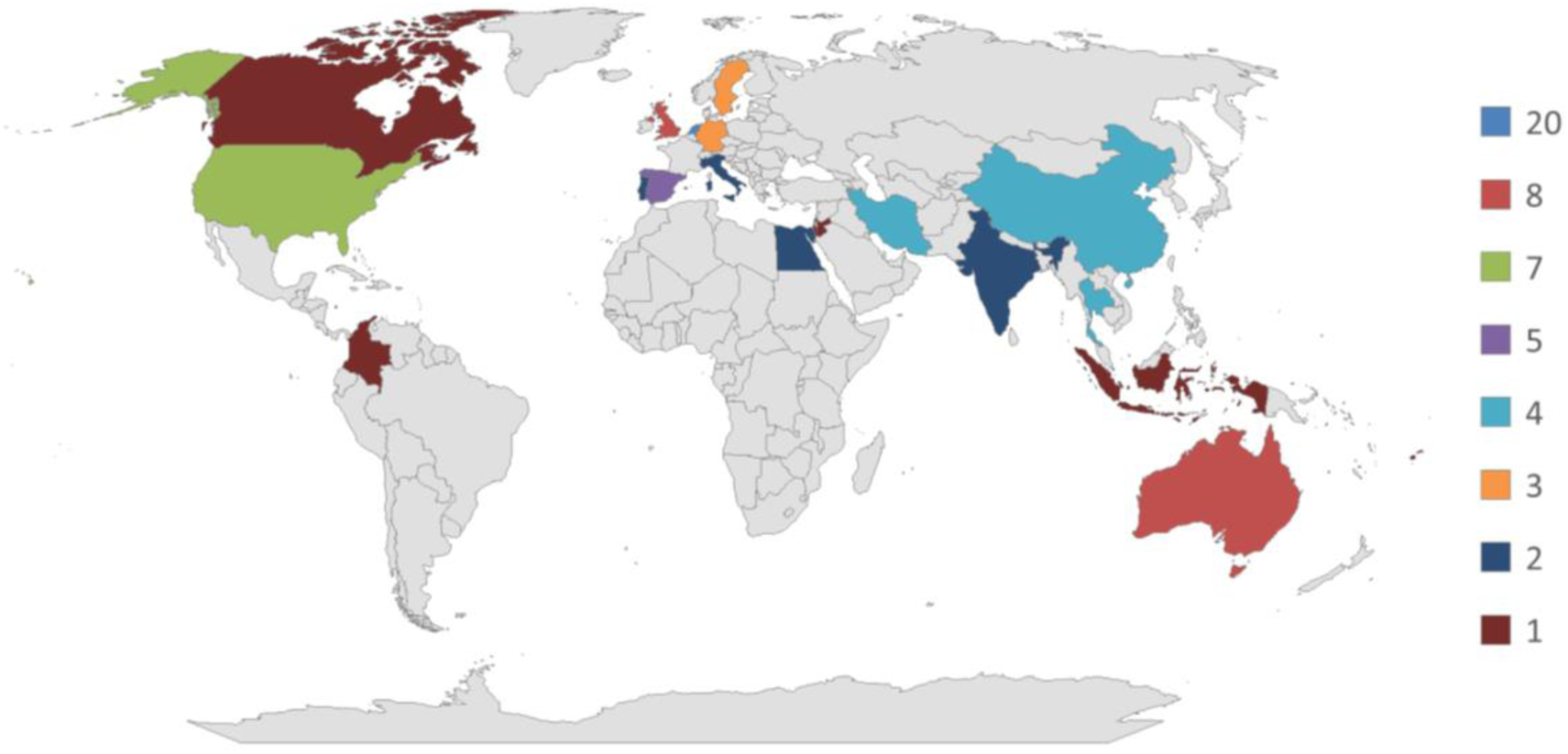
Countries by number of studies.

Figure 3 summarizes studies based on the study designs. The most common design was the cross-sectional study (n=20), capturing data on exposure and outcome at a single time point. This design was used predominantly for prevalence estimates and behavior assessments [14, 15, 31, 37, 39, 41, 43–45, 58, 61, 68, 69, 76, 80, 84, 87, 90, 96, 103]. Randomized controlled trials (RCTs) were used by 12 studies, offering randomization and control groups to test intervention efficacy [27, 38, 46, 47, 49, 56, 62, 66, 86, 93–95]. A total of 14 studies were observational, where outcomes were tracked without intervention, often in real-world or longitudinal settings [12, 28, 29, 32, 40, 42, 55, 59, 67, 70, 73, 81, 85, 89].

**Figure 3:**
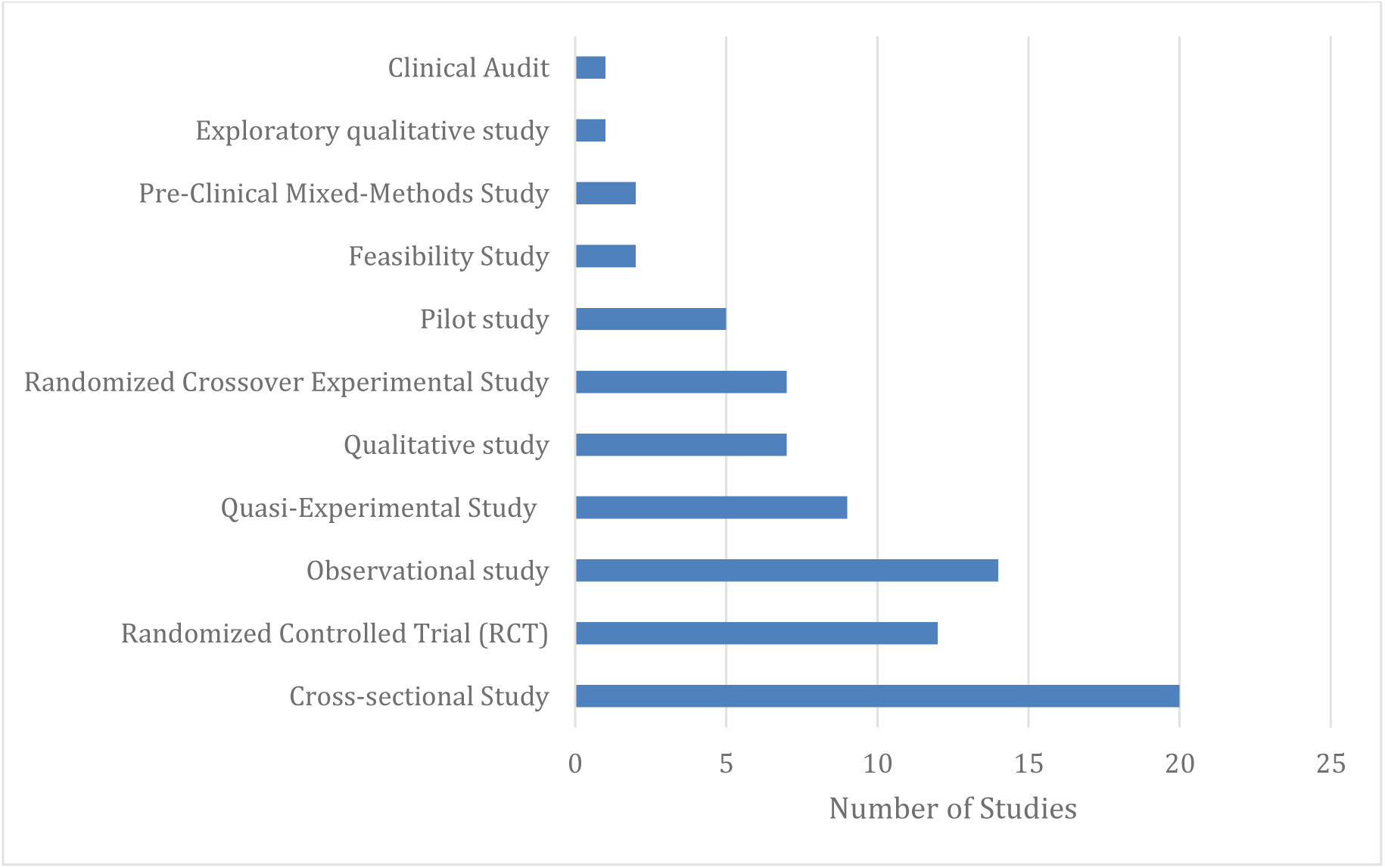
Number of studies by study design.

Quasi-experimental study designs were utilized by nine studies, providing controlled conditions without randomization and were typically used to assess device or footwear effects across time or conditions [48, 51, 60, 65, 72, 74, 77, 79, 98]. Seven studies utilized qualitative assessments methodologies such as interpretative phenomenological analysis and thematic coding to explore patient perspectives on footwear adherence, usability, and self-management [13, 36, 50, 75, 97, 100, 102].

A group of randomized crossover experimental studies (n=7) used within-subject designs with randomized order of conditions, allowing participants to serve as their own control. This study design was a common approach in biomechanics and gait research [33, 35, 53, 78, 82, 99, 101]. Smaller and preliminary investigations included pilot studies (n=5), which focused on early-stage feasibility and effect trends [34, 52, 71, 88, 91], and feasibility studies (n=2), which tested practical implementation without assessing outcomes [63, 64].

Two studies used a pre-clinical mixed-methods design, combining qualitative and quantitative methodologies data across iterative phases involving usability, validation, and device refinement [57, 92]. One exploratory qualitative study [83] focused on user experience in context, and one retrospective clinical audit [54] reviewed historical clinical data to assess practice patterns across expected standards.

The included studies represent a methodologically diverse body of evidence, spanning from exploratory, hypothesis-generating research to confirmatory randomized controlled trials. This diversity reflects the evolution of a maturing research field that increasingly integrates clinical effectiveness with user-centered design, implementation science, and contextually informed evaluation approaches.

The studies utilized a diverse set of outcomes to examine the effectiveness of the footwear interventions, as shown in Table 3. Among the studies investigating therapeutic footwear and insoles for people with diabetes, plantar pressure was the most common outcome, reported in 55 studies. Plantar pressure was assessed using peak plantar pressure (PPP) in 40 studies [29, 31, 33, 35, 37–39, 41–45, 47–49, 53, 55, 57, 58, 60, 63, 64, 69, 74, 77–79, 82, 84–86, 88, 90–92, 95, 96, 98, 99, 104], and pressure time integral (PTI) in 13 studies [37, 58, 60, 64, 69, 74, 77–79, 84, 85, 95, 104]. PPP evaluates the maximum pressure in the affected region while PTI quantifies the cumulative pressure applied over time to a specific area. Only one study reported more advanced plantar pressure metrics such as peak pressure gradient (PGrad), peak pressure time curve (PTC), peak pressure map (PMap), and peak pressure time map (PTM) [44], highlighting the limited adoption of these multidimensional approaches in routine clinical or research evaluation.

**Table 3:**
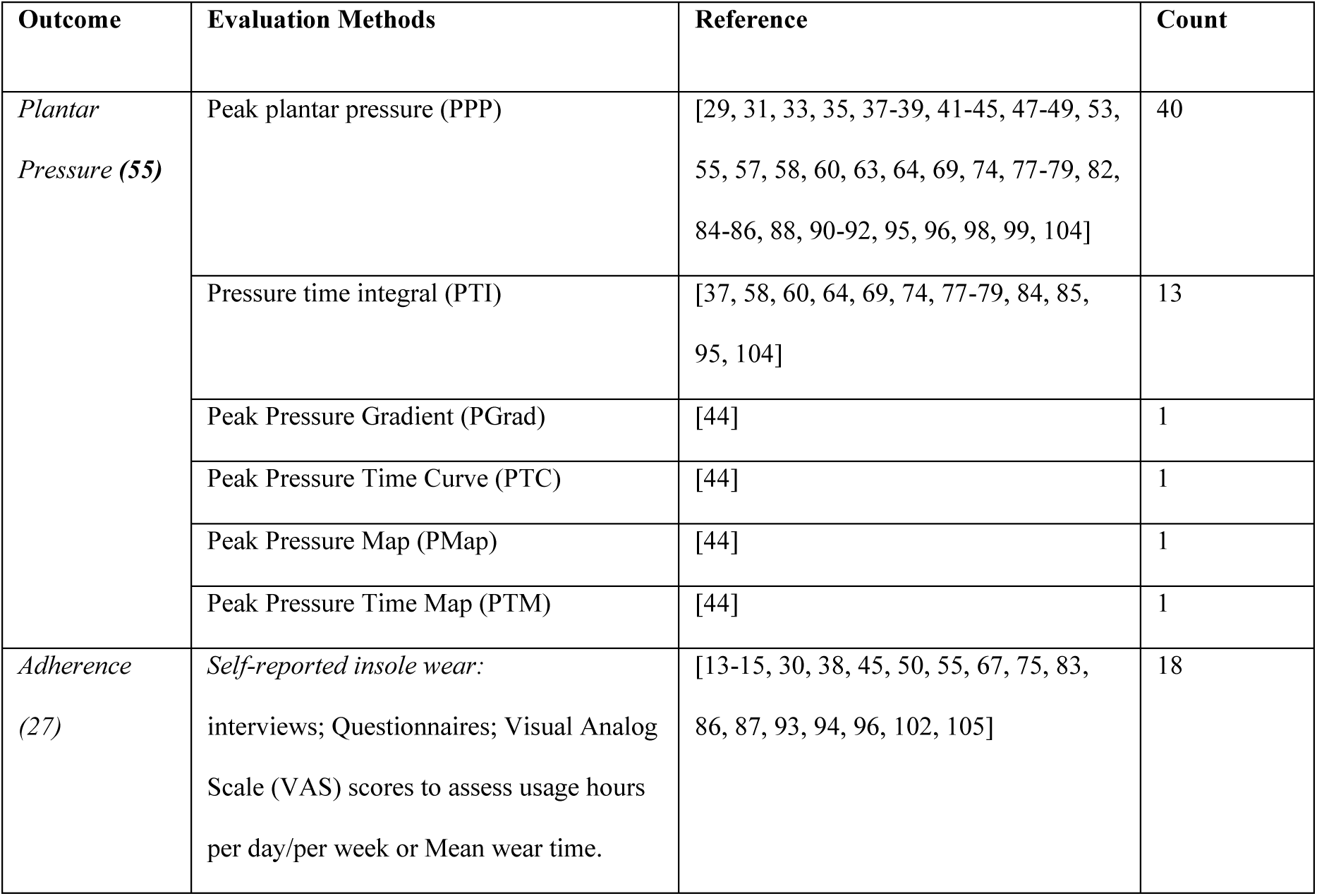
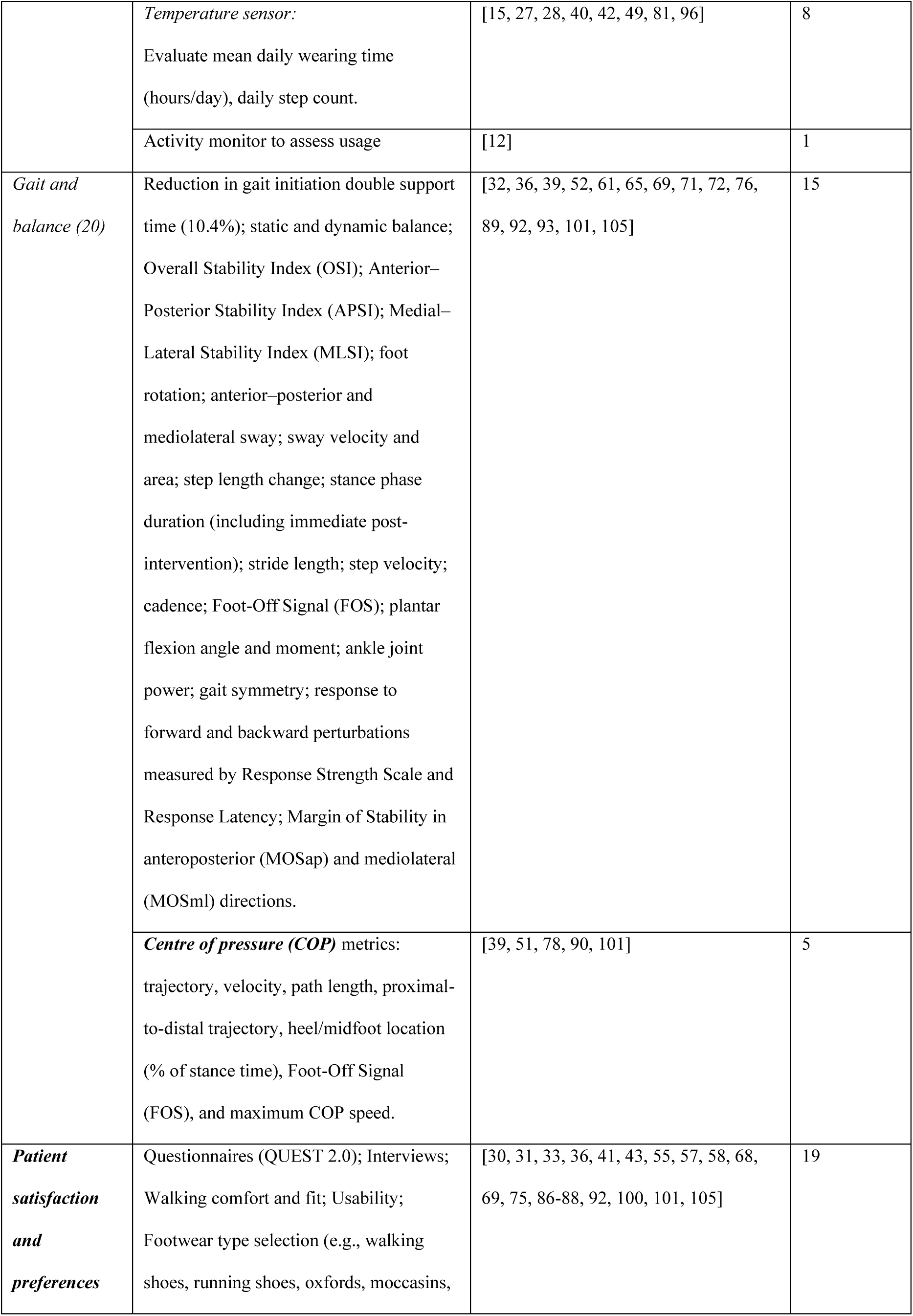
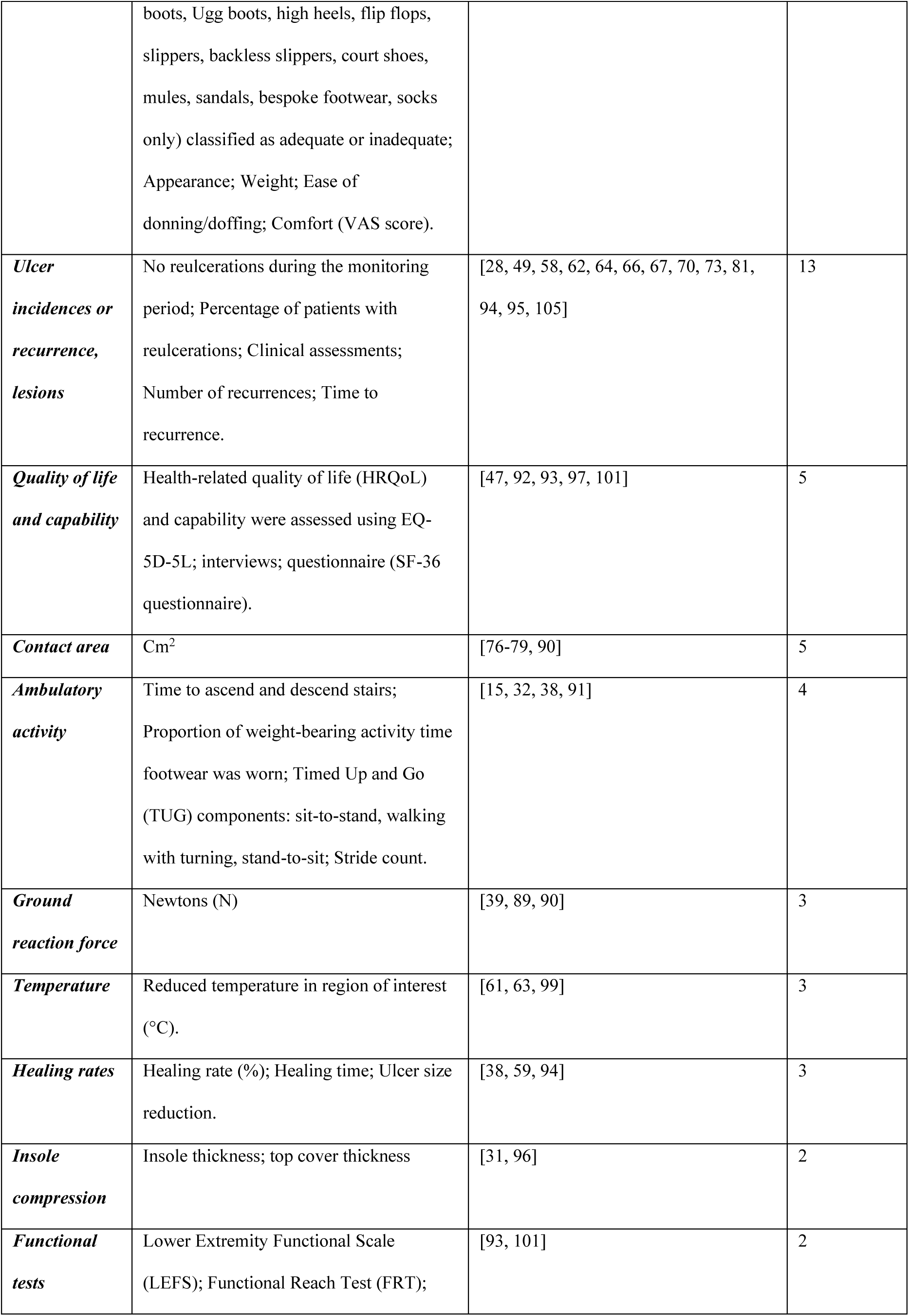
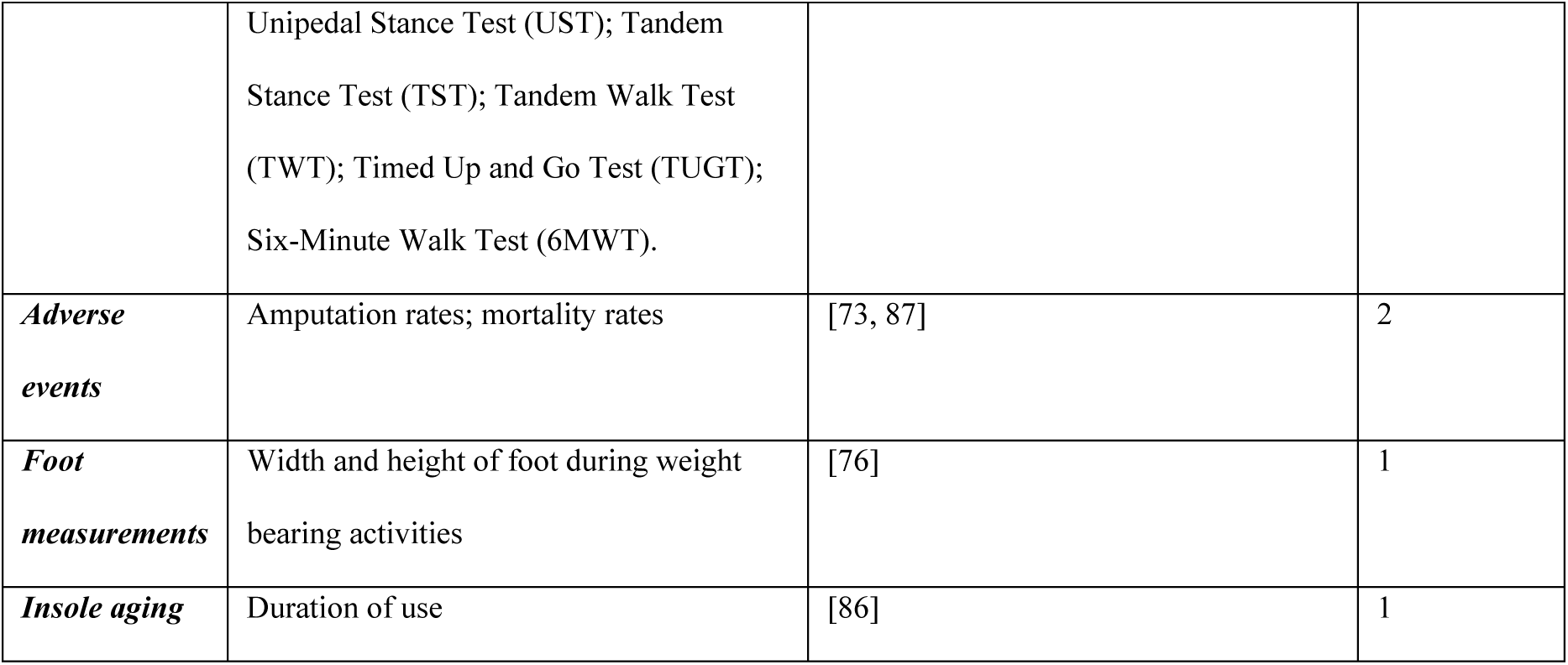
Studies by offloading Outcomes.

Adherence was the second most common outcome reported across the studies (n=27). Self-reported adherence was used in 18 studies [13–15, 30, 38, 45, 50, 55, 67, 75, 83, 86, 87, 93, 94, 96, 102, 105] through interviews, questionnaires, or visual analog scale (VAS) ratings. Objective monitoring techniques were applied in eight studies [15, 27, 28, 40, 42, 49, 81, 96], where temperature sensors were used to assess mean daily wear time and step count. In addition, one study [12] employed an activity monitor to capture physical activity data. Together, these measures reflect increasing efforts to quantify real-world device usage accurately.

Gait and balance were evaluated in 20 studies, with 15 studies analyzing gait and postural parameters such as double support time, stride length, sway velocity, and stability indices [32, 36, 39, 52, 61, 65, 69, 71, 72, 76, 89, 92, 93, 101, 105]. Additionally, center of pressure (COP) metrics was reported in five studies [39, 51, 78, 90, 101], assessing foot trajectory, velocity, and foot-off signals as markers of dynamic and static balance.

Patient satisfaction and preferences were reported in 19 studies using a combination of questionnaires like QUEST 2.0, structured interviews, and subjective assessments of fit, comfort, aesthetics, and ease of use [30, 31, 33, 36, 41, 43, 55, 57, 58, 68, 69, 75, 86–88, 92, 100, 101, 105]. Ulcer incidence or recurrence was reported in 13 studies [28, 49, 58, 62, 64, 66, 67, 70, 73, 81, 94, 95, 105], using clinical data to track reulceration frequency, time to recurrence, and number of affected patients.

Other outcomes were reported less frequently. Quality of life and capability were assessed in five studies [47, 92, 93, 97, 101] via validated tools such as EQ-5D-5L and SF-36. Contact area was measured in five studies [76–79, 90], and ambulatory activity was assessed in four studies [15, 32, 38, 91] through stair navigation time, Timed Up and Go (TUG), stride count and using an activity monitor [15, 32, 38, 91]. Ground reaction force [39, 89, 90], plantar temperature [61, 63, 99], and healing rates [38, 59, 94] were each reported in three studies. A small number of studies examined insole compression [31, 96], functional tests [93, 101], adverse events such as amputation or mortality [73, 87], foot measurements [76], and insole aging [86], each appearing in only 1–2 studies.

The use of diverse outcome measures suggests a prevailing research focus on biomechanical parameters, particularly PPP, while also demonstrating increasing attention to adherence monitoring, patient satisfaction, and clinical outcomes such as ulcer recurrence and healing. Notably, recent studies have incorporated adherence as a secondary outcome to evaluate the effectiveness of therapeutic footwear and insoles. The integration of both quantitative and subjective measures reflects a maturing research field with a growing emphasis on real-world effectiveness and user-centered care.

### Footwear Prescription Decision Factors

The footwear prescription decision factors are mapped using the World Health Organization’s (WHO) five dimensions of adherence: (1) patient-related, (2) condition-related, (3) therapy-related, (4) socioeconomic, and (5) healthcare system–related factors. The decision-making factors are summarized in Table 4, which consolidates variables cited in the eligibility criteria and those discussed in the results across the included studies.

**Table 4:**
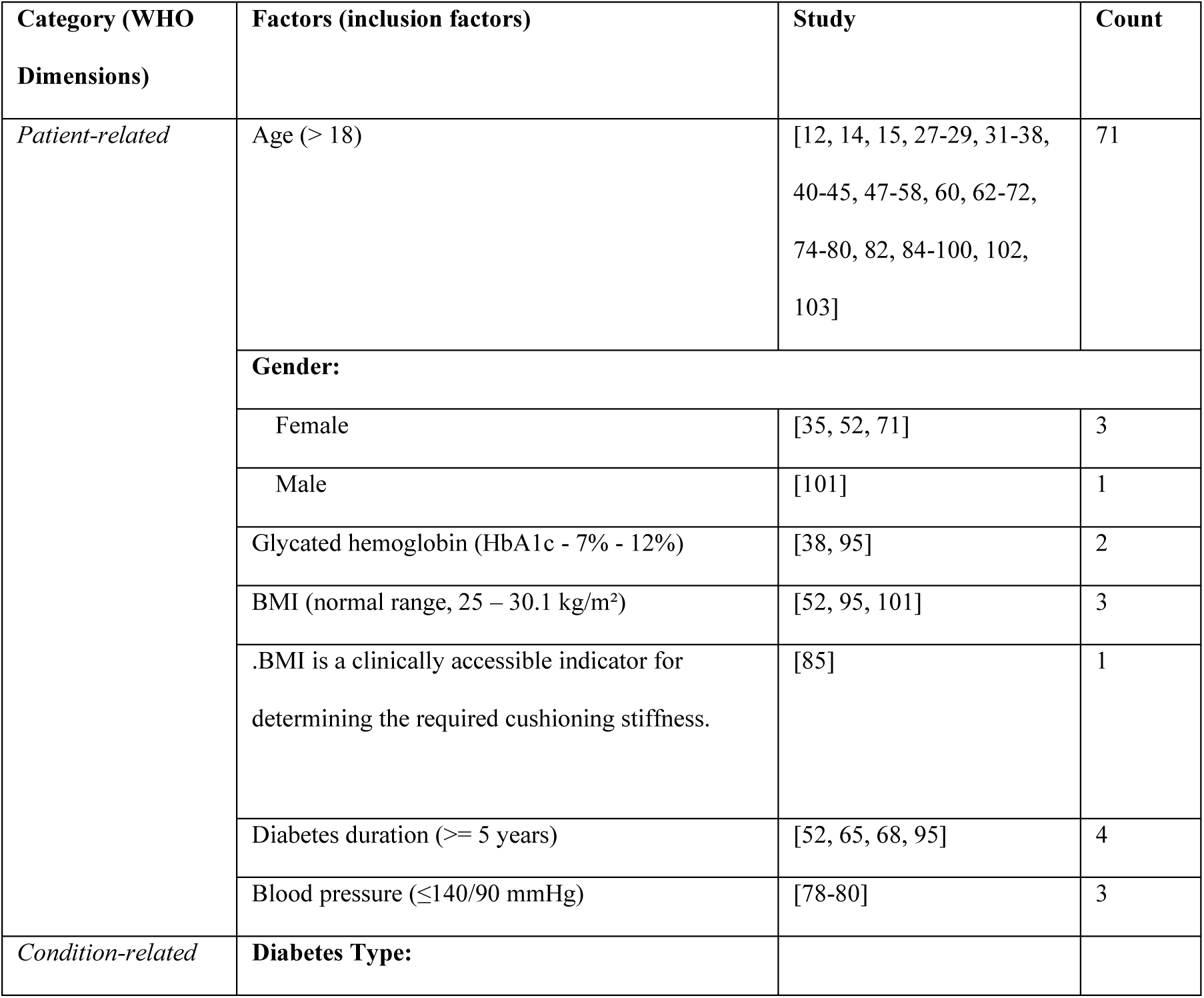
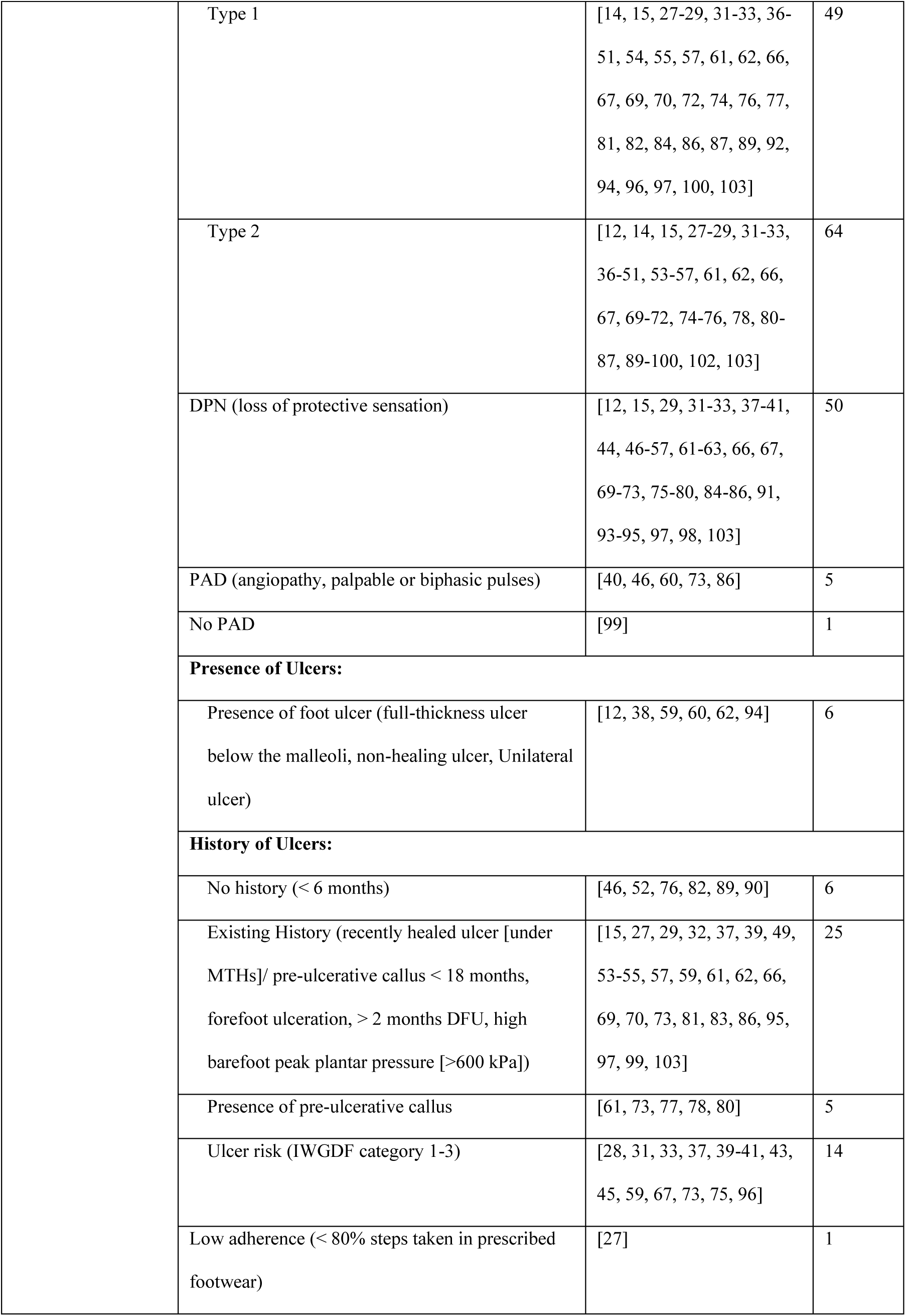
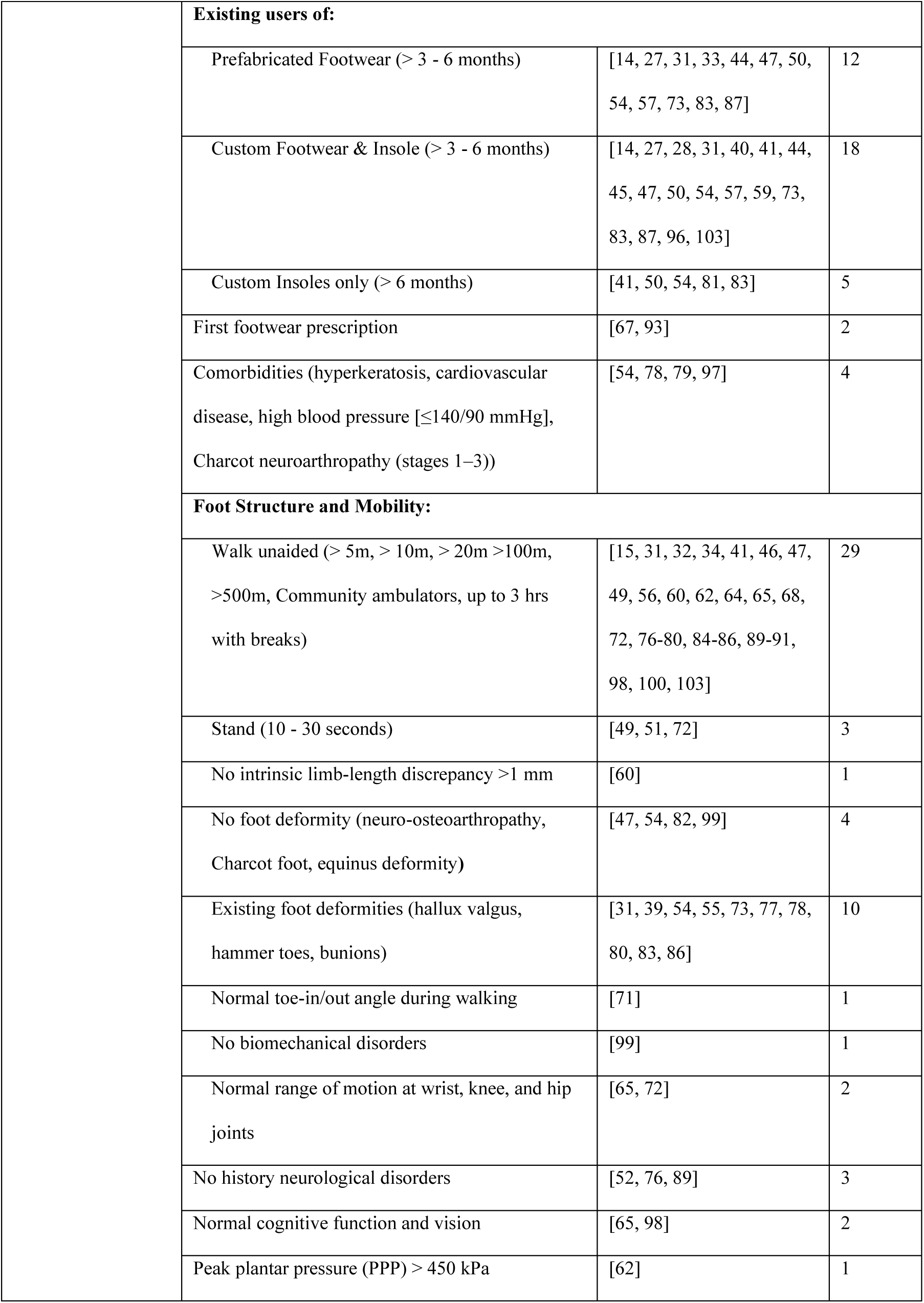
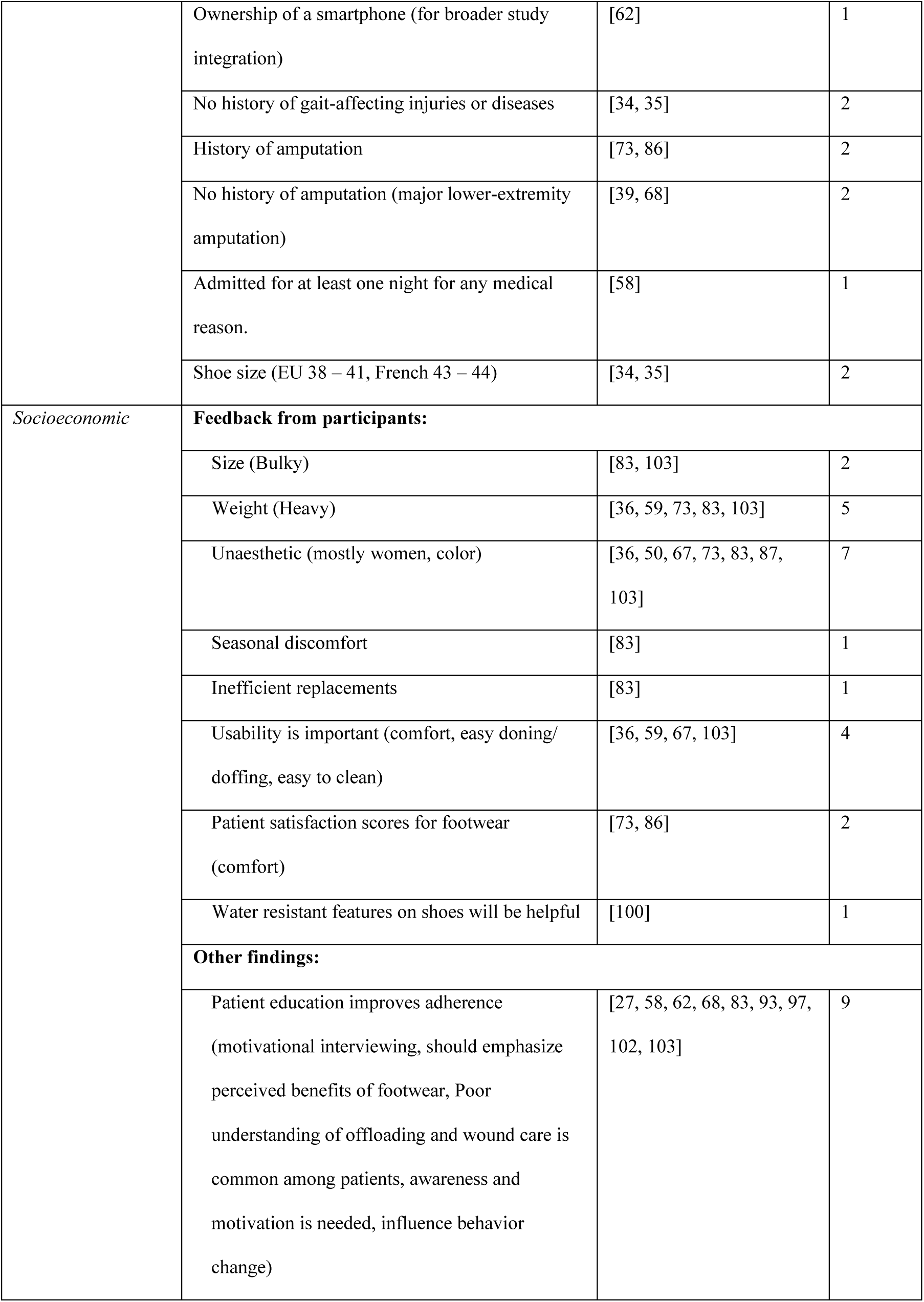
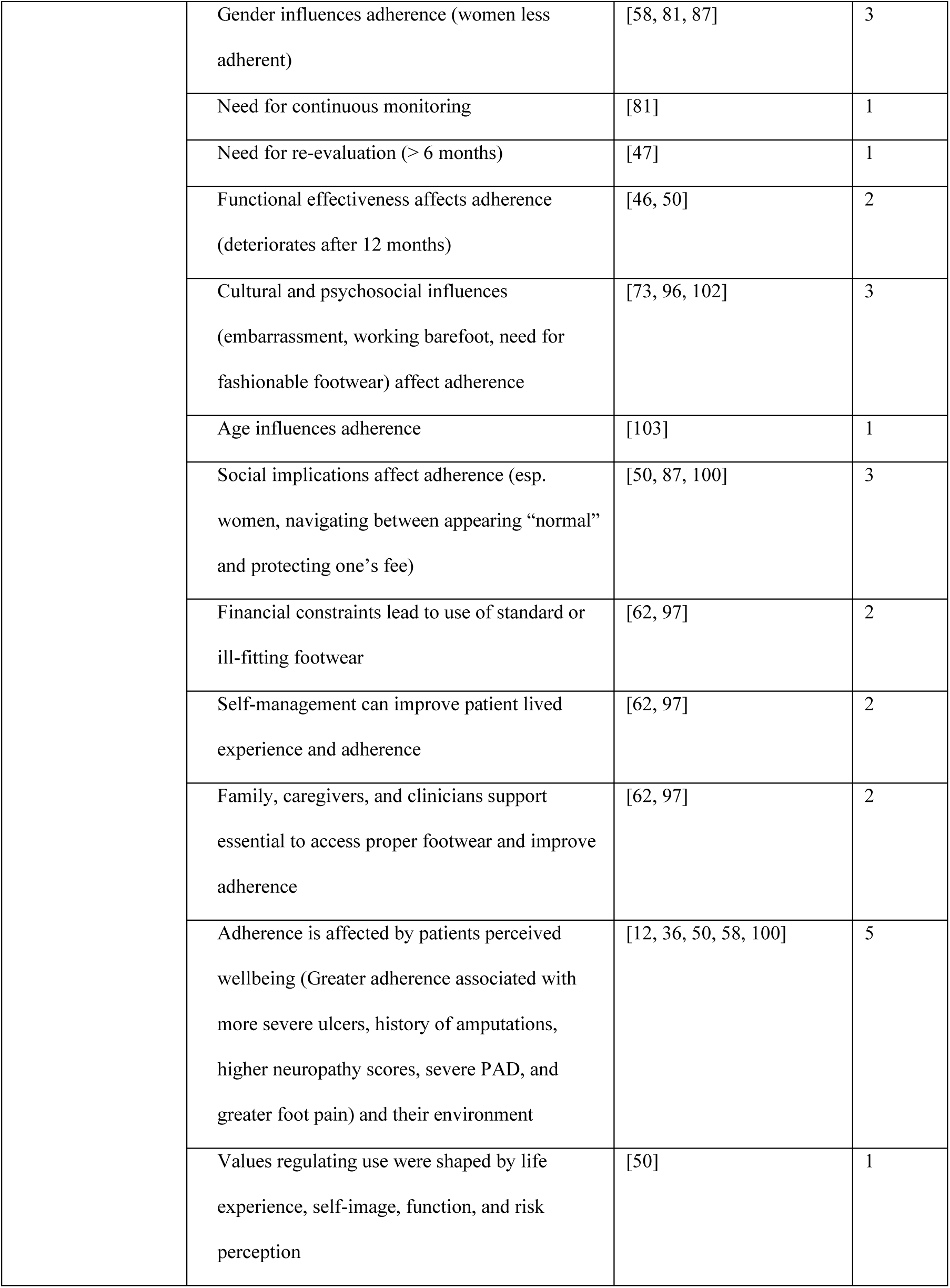
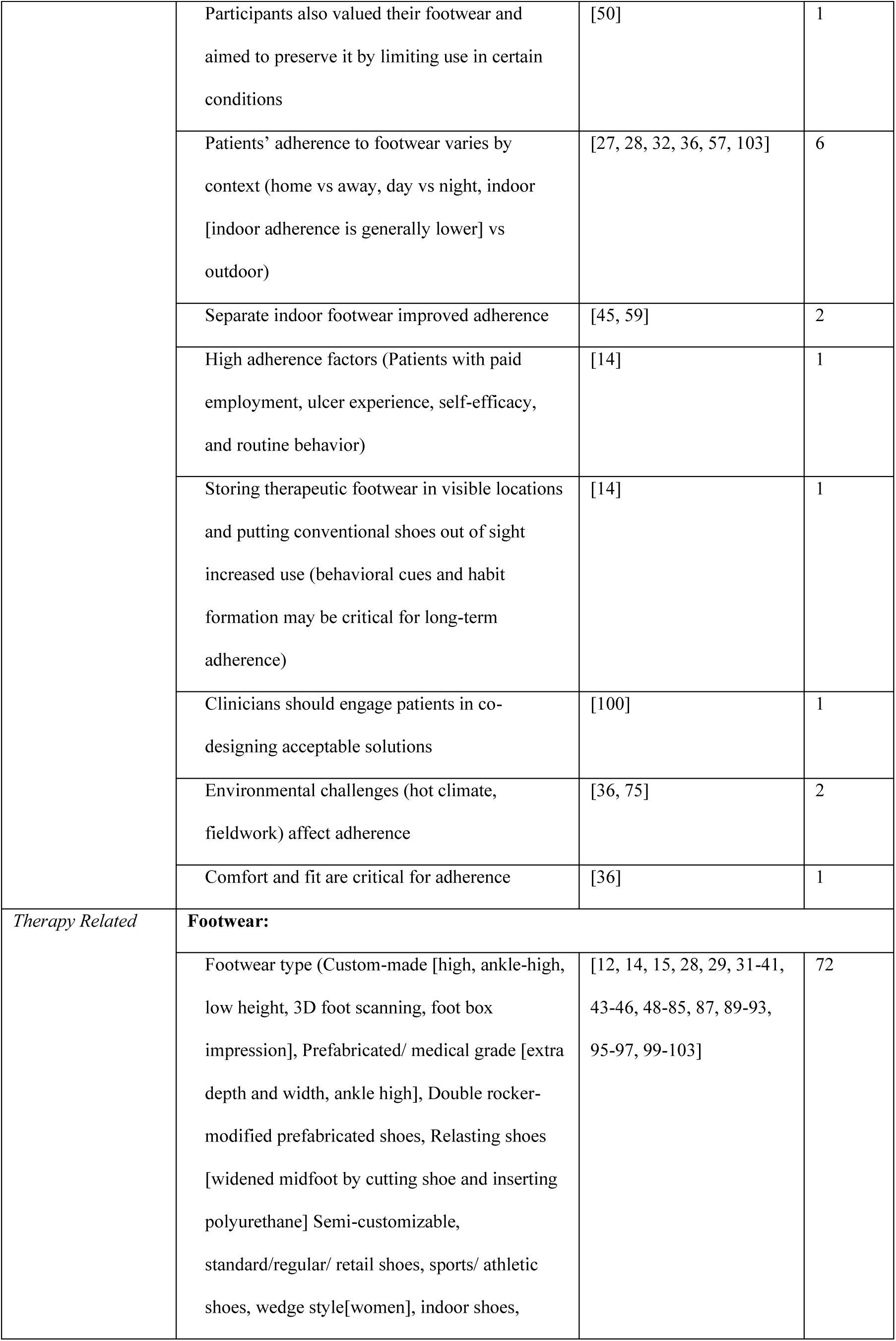
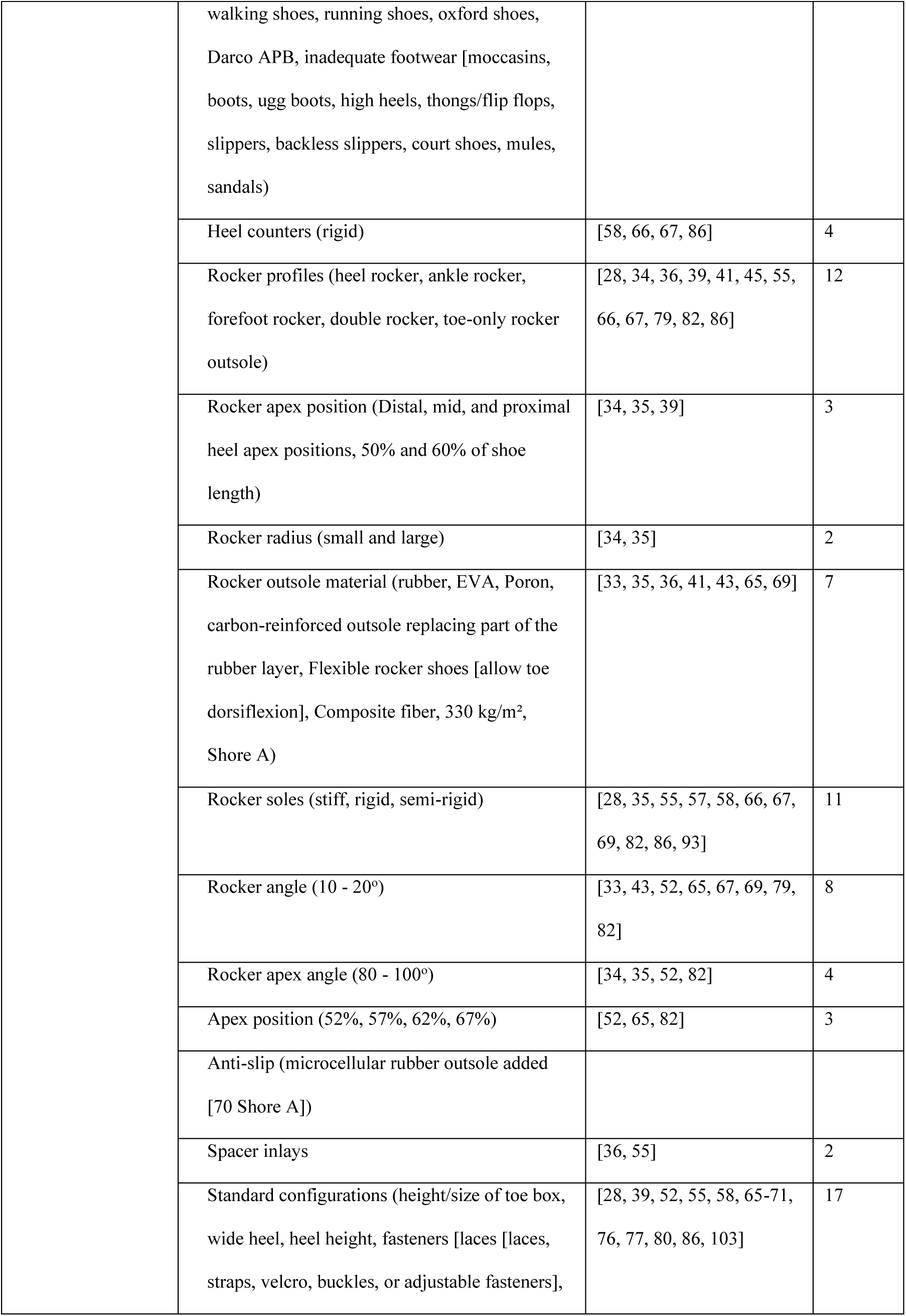
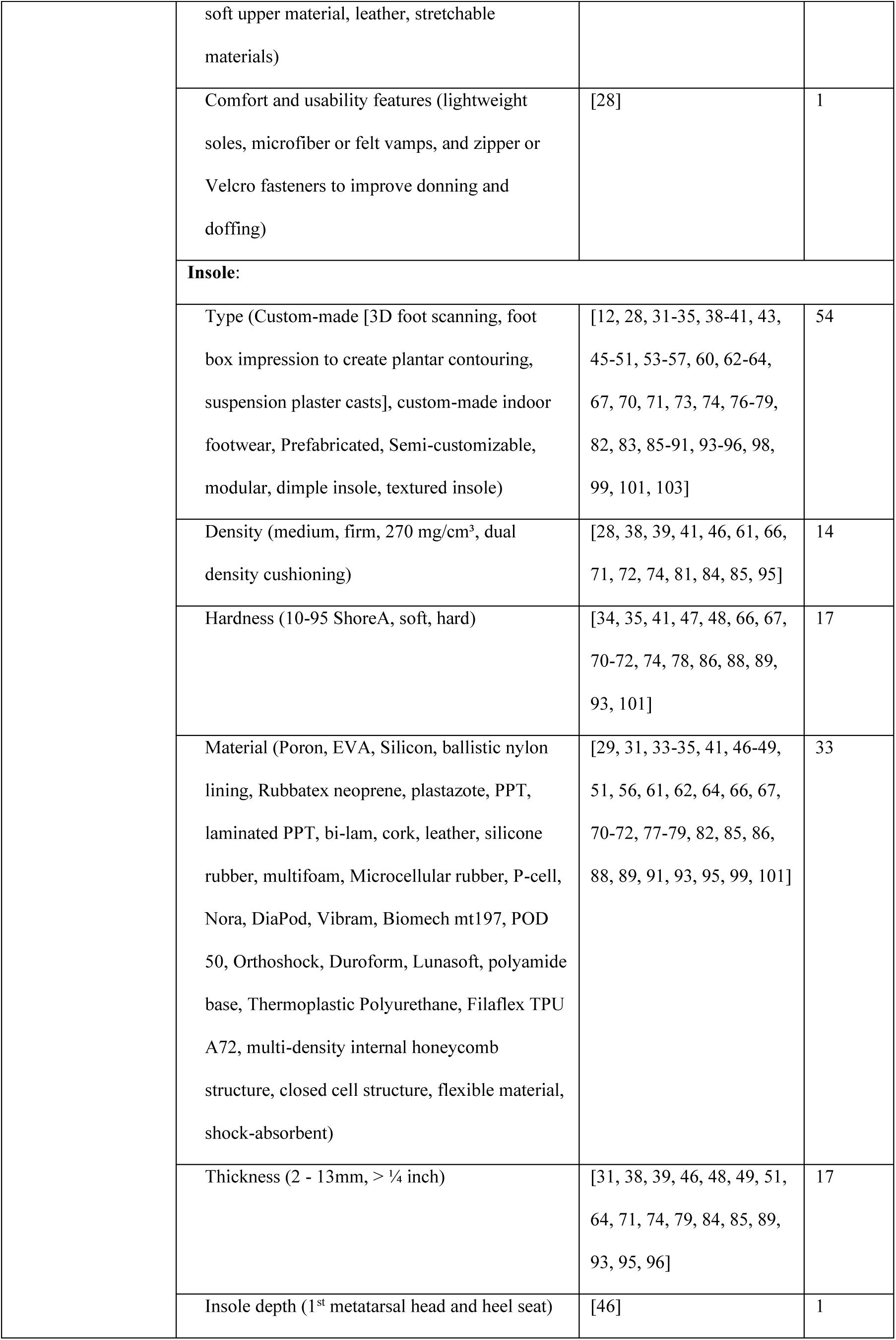
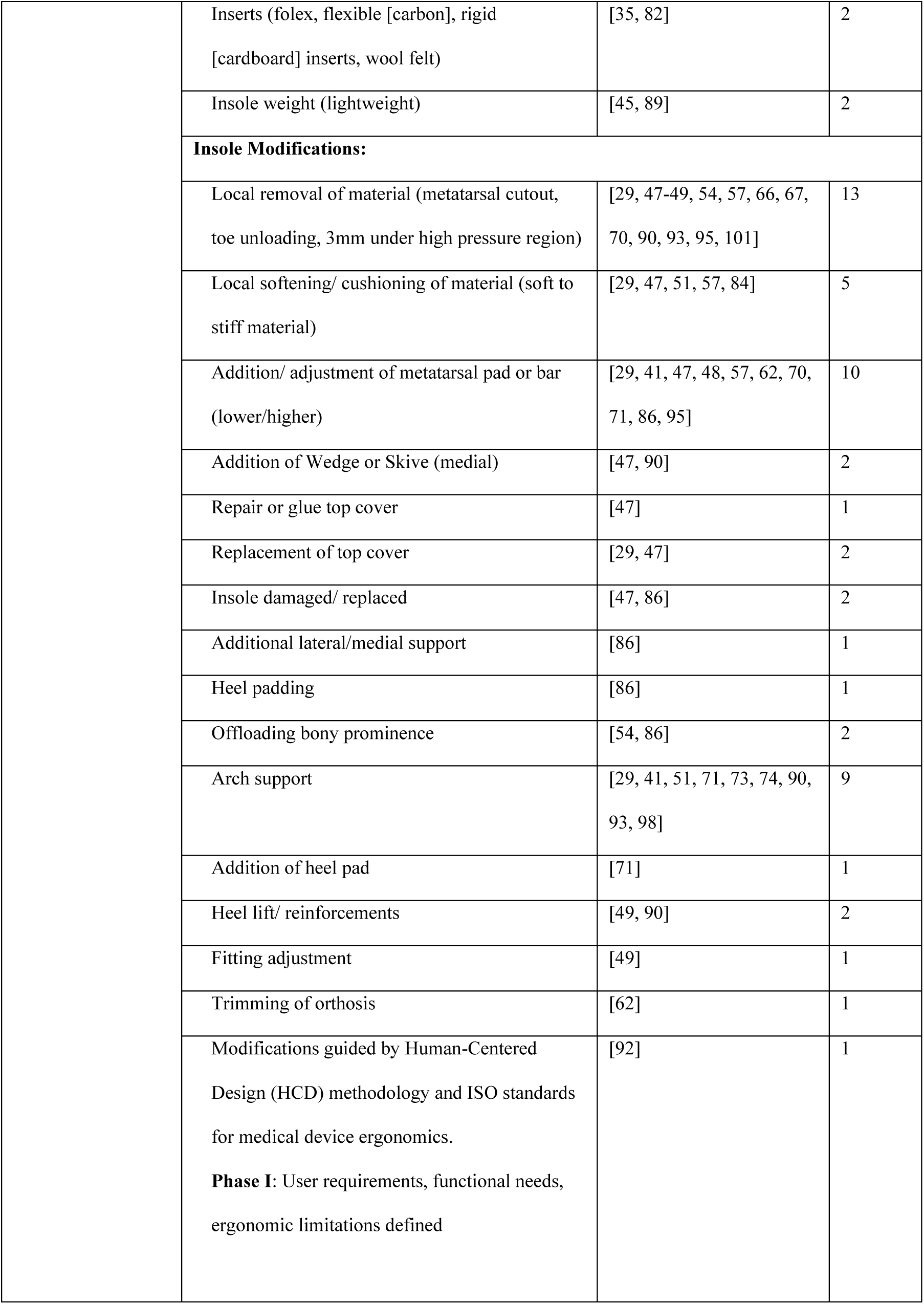
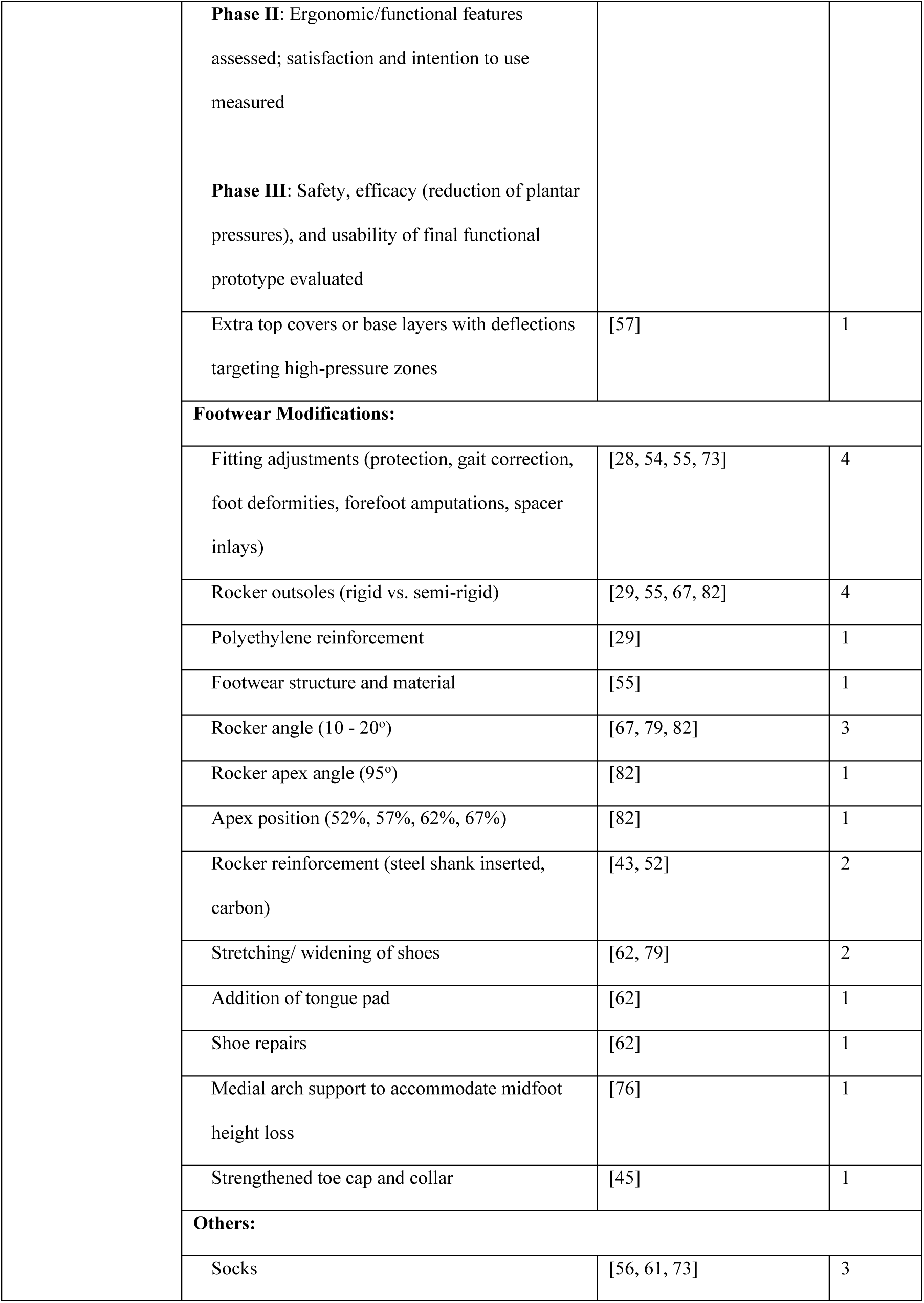
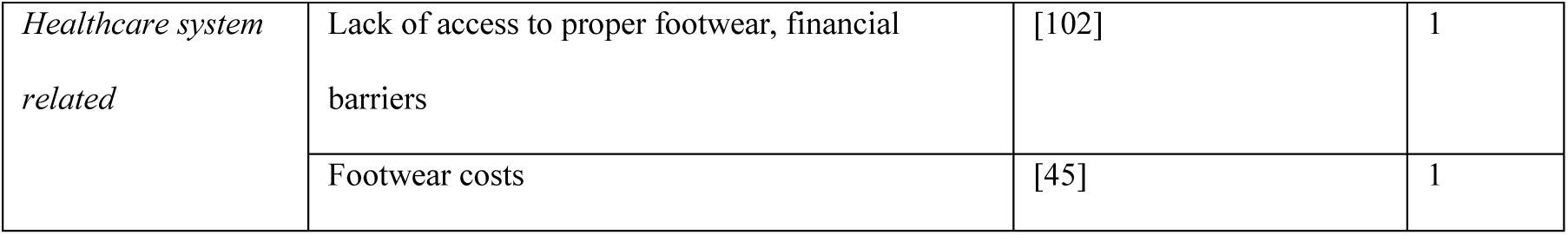
Footwear prescription decision factors.

#### Patient-Related Factors

Seventy-one (72) studies reported patient-related inclusion criteria. The most common factor was age, with all studies requiring participants aged >18 years [12, 14, 15, 27–29, 31–38, 40–45, 47–58, 60, 62–72, 74–80, 82, 84–100, 102, 103]. Gender-specific studies were rare, with only three studies focused on females [35, 52, 71] and one on males [101]. Clinical thresholds included HbA1c values between 7% and 12% (n=2) [38, 95], BMI between 25 and 30.1 kg/m² (n=3) [52, 95, 101], and systolic/diastolic blood pressure ≤140/90 mmHg (n=3) [78–80]. Four studies [52, 65, 68, 95] required diabetes duration ≥5 years, and one study used BMI to tailor insole cushioning density [85].

#### Condition-Related Factors

Included participants were diagnosed with either type 1 (n=49) [14, 15, 27–29, 31–33, 36–1, 54, 55, 57, 61, 62, 66, 67, 69, 70, 72, 74, 76, 77, 81, 82, 84, 86, 87, 89, 92, 94, 96, 97, 100, 103] or type 2 diabetes (n=64) [12, 14, 15, 27–29, 31–33, 36–51, 53–57, 61, 62, 66, 67, 69–72, 74–76, 78, 80–87, 89–100, 102, 103], with many studies involving both populations. DPN was reported across 50 studies [12, 15, 29, 31–33, 37–41, 44, 46–57, 61–63, 66, 67, 69–73, 75–80, 84–86, 91, 93–95, 97, 98, 103], while peripheral arterial disease (PAD) was reported across 5 studies [40, 46, 60, 73, 86]. One study specifically excluded participants with PAD [99].

Foot risk status and ulcer history were important inclusion markers. Six studies recruited participants with current ulcers [12, 38, 59, 60, 62, 94]; 25 studies included those with recent or healed ulcers within the previous 18 months [15, 27, 29, 32, 37, 39, 49, 53–55, 57, 59, 61, 62, 66, 69, 70, 73, 81, 83, 86, 95, 97, 99, 103]. Conversely, six studies [46, 52, 76, 82, 89, 90] required no recent ulceration. Presence of pre-ulcerative callus was used as inclusion criteria in five studies [61, 73, 77, 78, 80], and 14 studies [28, 31, 33, 37, 39–41, 43, 45, 59, 67, 73, 75, 96] included patients with stratified risk using the IWGDF ulcer risk categories 1 to 3.

Footwear usage history was another common factor for inclusion. Prefabricated footwear use (>3–6 months) was used in 12 studies [14, 27, 31, 33, 44, 47, 50, 54, 57, 73, 83, 87], while 18 studies included participants already using custom-made footwear and insoles [14, 27, 28, 31, 40, 41, 44, 45, 47, 50, 54, 57, 59, 73, 83, 87, 96, 103]. Five studies included patients who used custom insole use alone [41, 50, 54, 81, 83]. While most of the studies only included patients who were existing users of footwear or insole, only two studies enrolled participants receiving footwear for the first time [67, 93]. Additionally, one study included existing users of footwear with documented low adherence [27].

Comorbidities such as cardiovascular disease, controlled hypertension, Charcot neuroarthropathy, and hyperkeratosis were used to select participants in four studies[54, 78, 79, 97]. Functional mobility was emphasized in 29 studies, which required unaided walking across distances ranging from five meters to greater than 500 meters [15, 31, 32, 34, 41, 46, 47, 49, 56, 60, 62, 64, 65, 68, 72, 76–80, 84–86, 89–91, 98, 100, 103]. Standing unaided (10–30 seconds) was an inclusion criterion in three studies[49, 51, 72]. Some studies also required participants to be free from gait-affecting injuries or diseases (n=2) [34, 35], major neurological disorders (n=3) [52, 76, 89], and limb-length discrepancies (n=1) [60]. Additional criteria included smartphone ownership (n=1) [62], recent hospital admission (n=1) [58], and shoe size within predefined ranges (n=2) [34, 35].

Foot structure and motor capacity were used for inclusion. Four studies excluded individuals with deformities [47, 54, 82, 99], while ten studies included patients with mild deformities [31, 39, 54, 55, 73, 77, 78, 80, 83, 86]. Joint range of motion was assessed in two studies, toe-in/out angle in one [71], and peak plantar pressure thresholds above 450 kPa in another [62]. Cognitive and visual competence were included in two studies to ensure informed participation [65, 98]. Amputation status was variably used to include participants across studies, as two studies included patients with a history of amputation [73, 86] and two excluded them [39, 68].

#### Therapy-Related Factors

Therapy-related factors were reported in 72 studies. A variety of footwear types were described, including custom-made, prefabricated, modular, wedge, retail, sports, and indoor shoes. Some of these studies examined open footwear styles such as mules or thongs were described in a subset of studies. Several footwear configurations were considered across studies, with rocker soles being the most common feature considered before prescription. Rocker profiles were considered in 12 studies with variations in forefoot, heel, and double rocker configurations [28, 34, 36, 39, 41, 45, 55, 66, 67, 79, 82, 86]. Common features of rockers considered included adjustments to rocker angles by eight studies [33, 43, 52, 65, 67, 69, 79, 82], apex positions (n=3), and apex angles in four studies [34, 35, 52, 82], outsole materials [33, 35, 36, 41, 43, 65, 69], rocker radii by two studies [34, 35], density by 11 studies [28, 35, 55, 57, 58, 66, 67, 69, 82, 86, 93].

Other footwear attributes included rigid heel counters (n=4) [58, 66, 67, 86], wide toe boxes, soft uppers, fastenings options, lightweight or water-resistant design options discussed by 17 studies [28, 39, 52, 55, 58, 65–71, 76, 77, 80, 86, 103], anti-slip outsoles, and spacer inlays in two studies [36, 55]. One study specifically highlighted comfort and usability features of the footwear [28].

While footwear modifications are a common aspect of footwear prescription, few studies reported the type of modifications. Fittings adjustments are common, where footwear is adjusted at initial fitting [28, 54, 55, 73]. Rocker profile medication was also seen across studies, with four studies modifying rocker outsoles [29, 55, 67, 82], three studies on rocker angles [67, 79, 82], two studies adding rocker reinforcement [43, 52] and one study modifying rocker apex angle and apex position [82]. Other footwear modifications included stretching or widening of shoes [62, 79], modifying tongue pads and performing shoe repairs [62], medial arc support [76], and reinforcing toe cap and collar [45].

Fifty-four studies reported insole characteristics as part of the results. This included type of insole such as custom-made, prefabricated, semi-custom, modular, and textured insoles. Insole density was reported by 14 studies [28, 38, 39, 41, 46, 61, 66, 71, 72, 74, 81, 84, 85, 95], and hardness in 17 studies [34, 35, 41, 47, 48, 66, 67, 70–72, 74, 78, 86, 88, 89, 93, 101]. Materials were reported in 33 studies [29, 31, 33–35, 41, 46–49, 51, 56, 61, 62, 64, 66, 67, 70–72, 77–79, 82, 85, 86, 88, 89, 91, 93, 95, 99, 101], and insole thickness in 17 studies [31, 38, 39, 46, 48, 49, 51, 64, 71, 74, 79, 84, 85, 89, 93, 95, 96]. Less common insole features reported included weight in two studies [45, 89], inserts in two studies [35, 82], and depth in one study [46].

Insole modifications were more common across studies in comparison to footwear modifications. Thirteen studies described localized material removal [29, 47–49, 54, 57, 66, 67, 70, 90, 93, 95, 101], and five reported softening or cushioning strategies [29, 47, 51, 57, 84]. Addition of metatarsal bars and pads were a common modification utilized across ten studies [29, 41, 47, 48, 57, 62, 70, 71, 86, 95]. Another common modification was addition of arch support reported in nine studies [29, 41, 51, 71, 73, 74, 90, 93, 98]. Additional modifications included heel lifts [49, 90], addition of wedges [47, 90], repair or replacement of top cover [29, 47], and providing replacement insoles [47, 86]. A few studies focused modifications on additional support and heel padding [86], offloading bony prominence [54, 86], addition of heel pad [71], heel lift or reinforcements [49, 90], fitting adjustments [49], and trimming of orthosis [62]. Few studies reported use of extra top covers with deflections [57] and medial arch supports [76]. One study conducted structured modifications guided by Human-Centered Design (HCD) methodology and ISO standards for medical device ergonomics [92].

#### Socioeconomic Factors

Socioeconomic factors were commonly discussed among footwear adherence studies, highlighting factors behind low adherence. Based on patient feedback, adherence is affected by footwear size [83, 103], weight [36, 59, 73, 83, 103], appearance [36, 50, 67, 73, 83, 87, 103], seasonal discomfort and inefficient replacements [83], usability [36, 59, 67, 103], comfort [73, 86], and additional features such as water resistance [100].

Other studies reported that patient education is associated with better adherence [27, 58, 62, 68, 83, 93, 97, 102, 103]. Gender [58, 81, 87], age [103], and social context [50, 87, 100] affect adherence behavior, with lower adherence observed among women. Context-dependent use [27, 28, 32, 36, 57, 103] has an impact on adherence with studies demonstrating that adherence is generally lower at home or at night. Cultural and psychosocial factors such as embarrassment, fashion preferences, and work routines also contribute to nonadherence [73, 96, 102].

Clinical factors such as disease severity affect adherence, with greater adherence linked to prior ulceration, pain, neuropathy, and amputations [12, 36, 50, 58, 100]. Comfort and fit [36], and the functional effectiveness of footwear [46, 50] over time are also essential, while environmental conditions [36, 75] and financial constraints [62, 97] further affect consistent use of footwear.

Monitoring and behavioral strategies such as continuous monitoring [81] and evaluation [47], habitual preferences such as storing shoes visibly [14], patients with paid employment self-efficacy, and routine behavior have shown potential in improving adherence. Social support from family, caregivers, and clinicians [62, 97] is also critical, and involving patients in co-designing acceptable footwear solutions may enhance uptake and long-term use [100].

#### Healthcare System–Related Factors

Two studies reported barriers related to access and cost. Participants identified limited availability of appropriate therapeutic footwear [102] and footwear costs [45] as barriers to access of care.

### Mapping to FHIR

Table 5 presents a structured framework for creating a dataset suitable for training AI and machine learning models. It categorizes key decision-making factors into input and output variables, aligning each with FHIR resources, coding systems, data types, units, and sample values. Input variables include patient-related data, condition-specific indicators, socioeconomic factors, and healthcare-related elements. These variables are primarily Boolean, integer, decimal, or coded, facilitating straightforward integration into binary and categorical models.

**Table 5:**
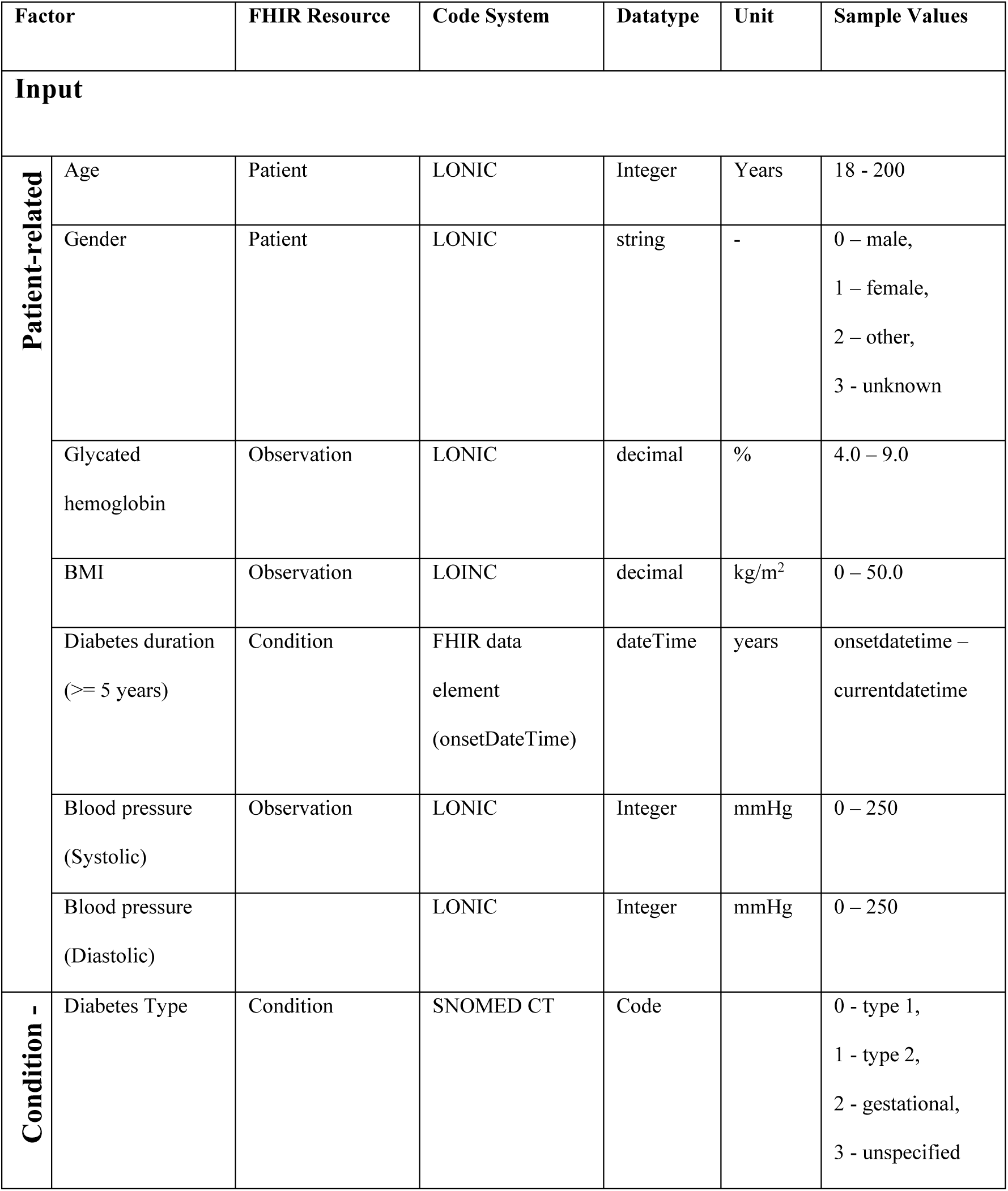
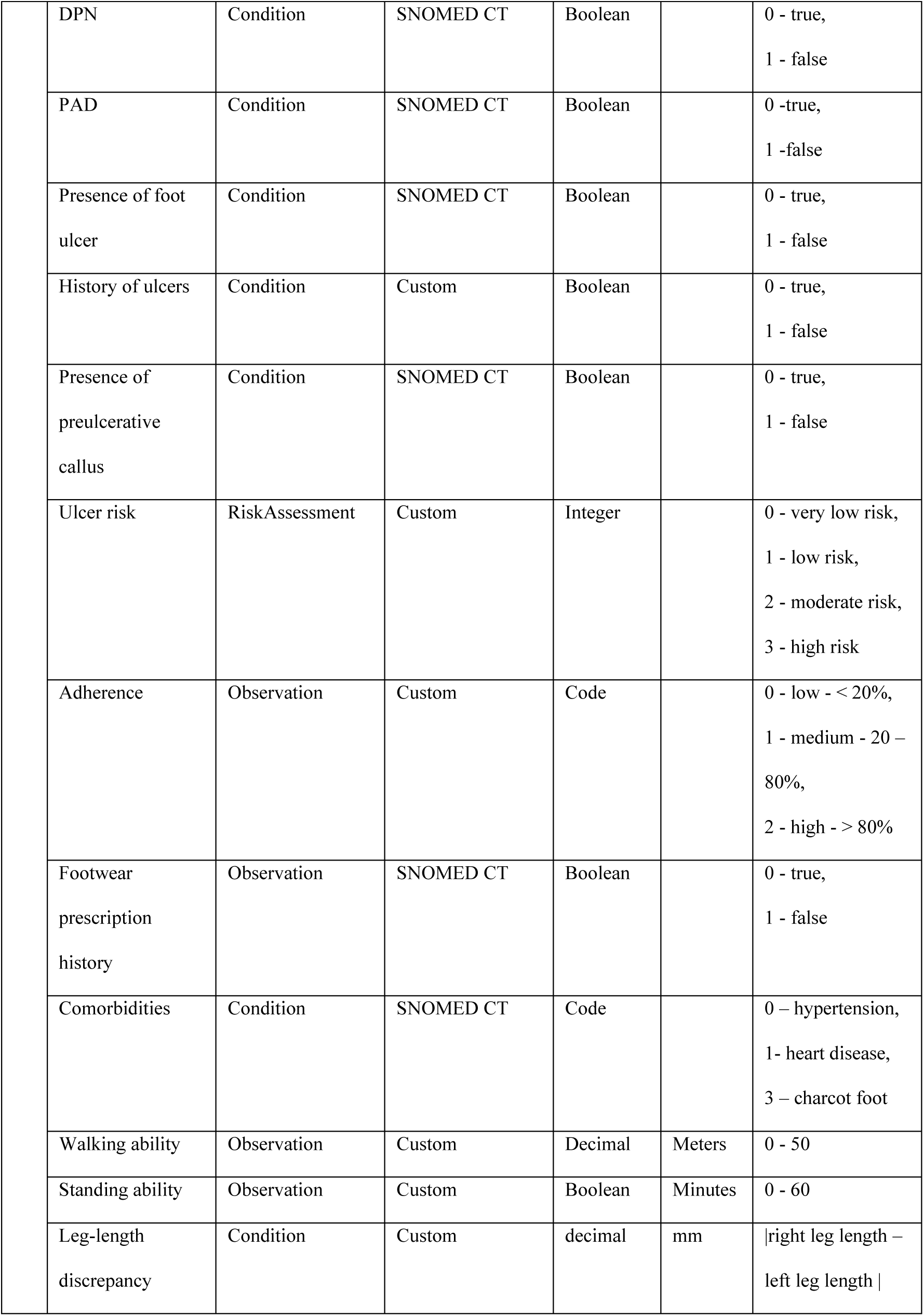
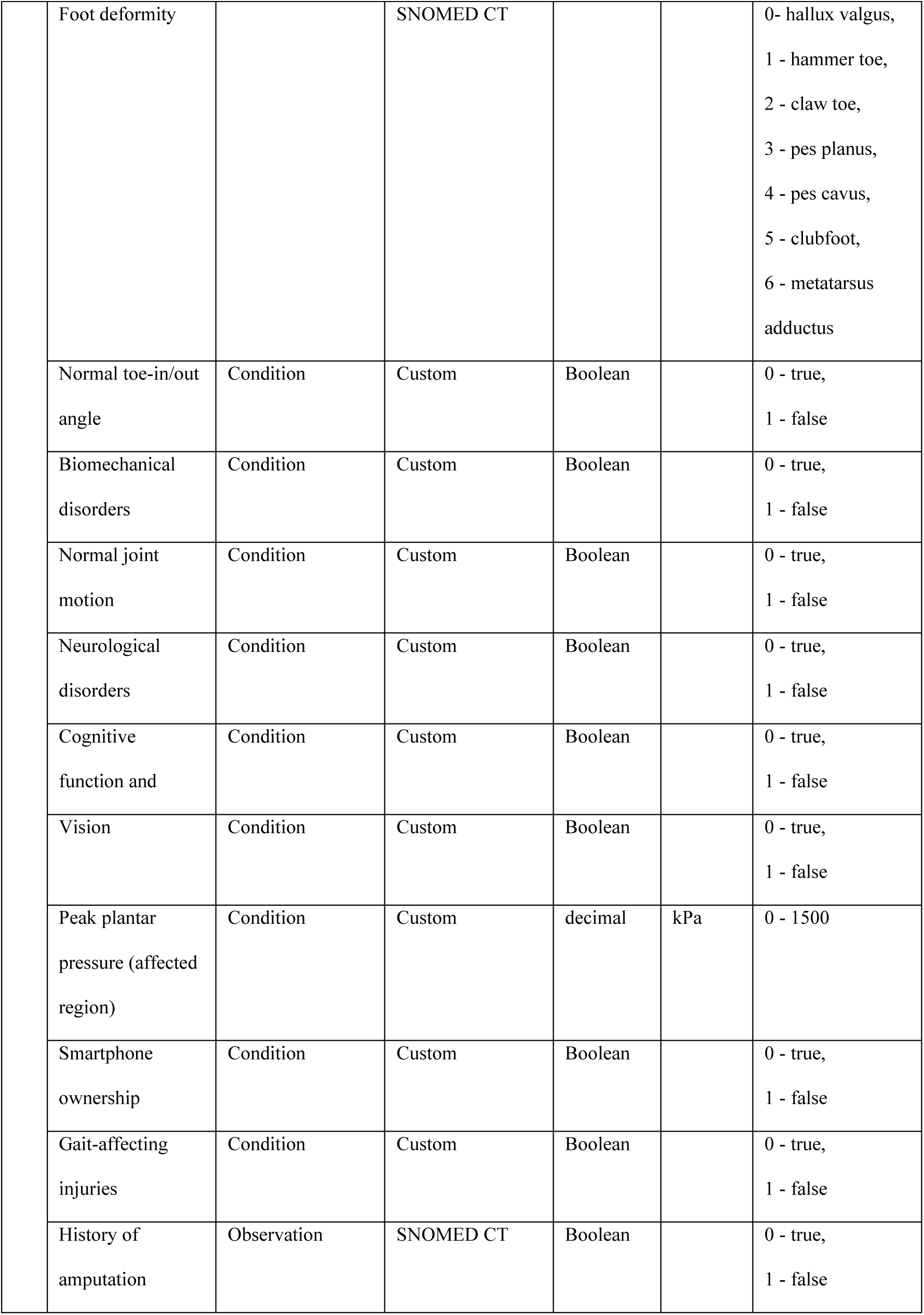
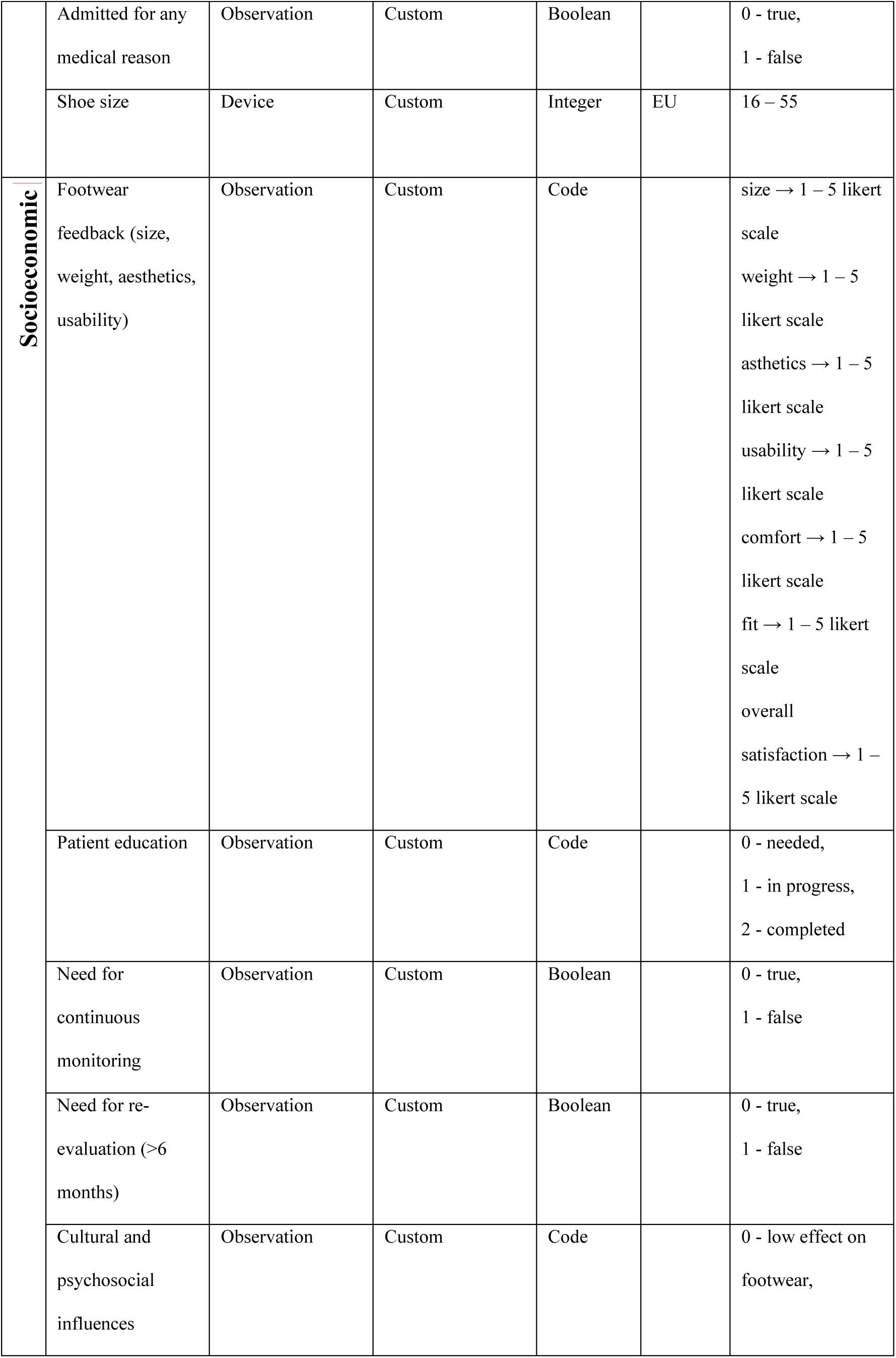
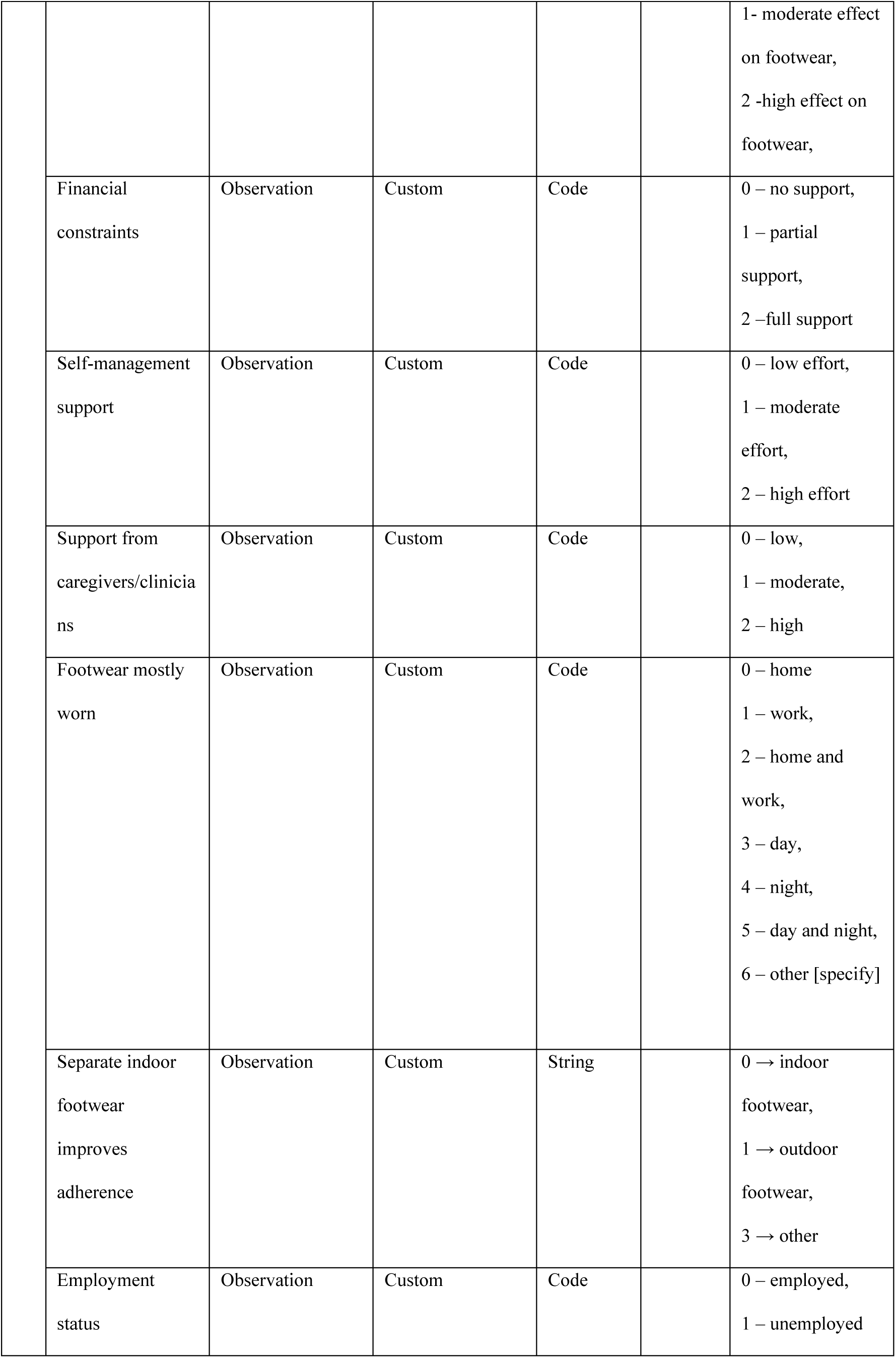
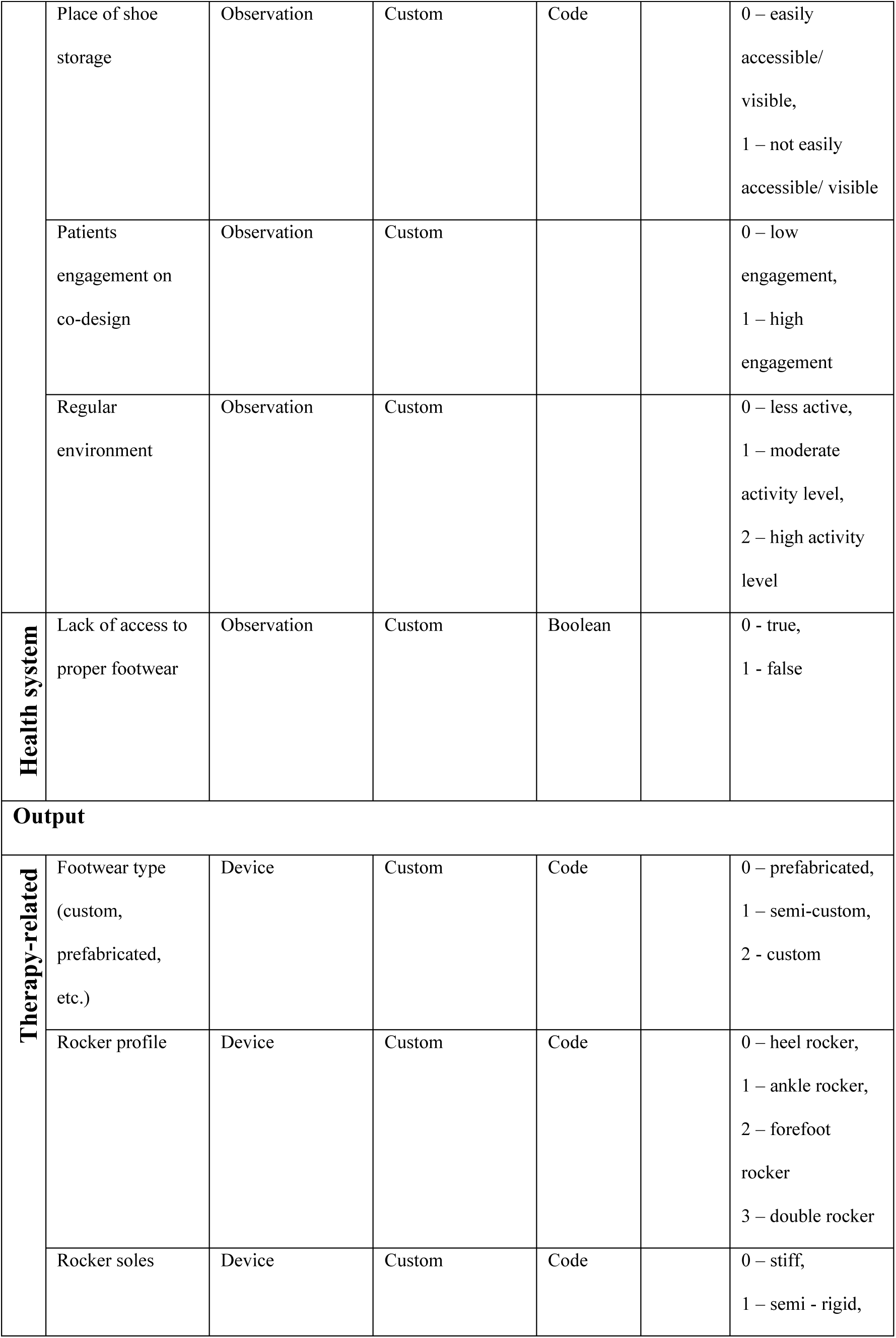
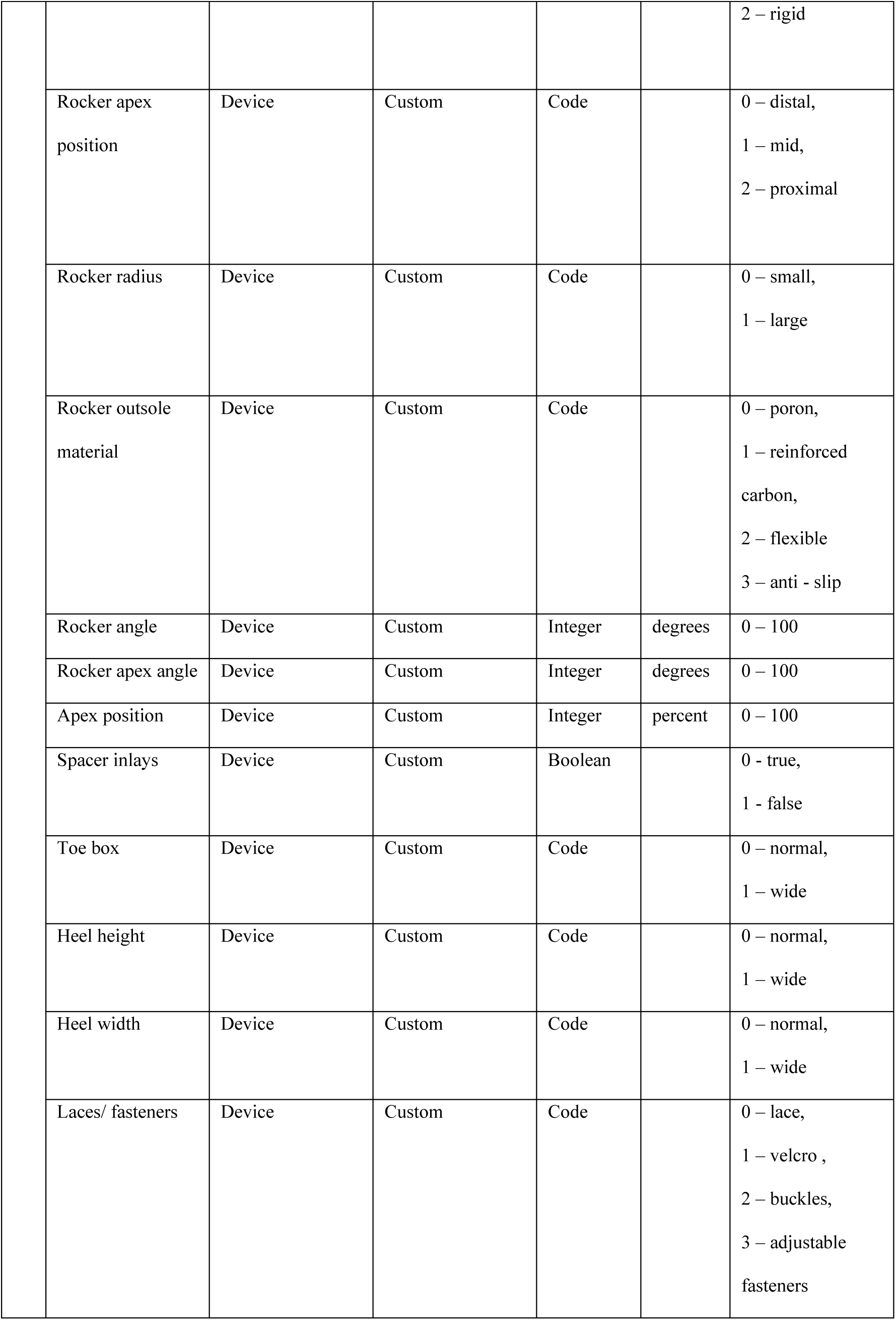
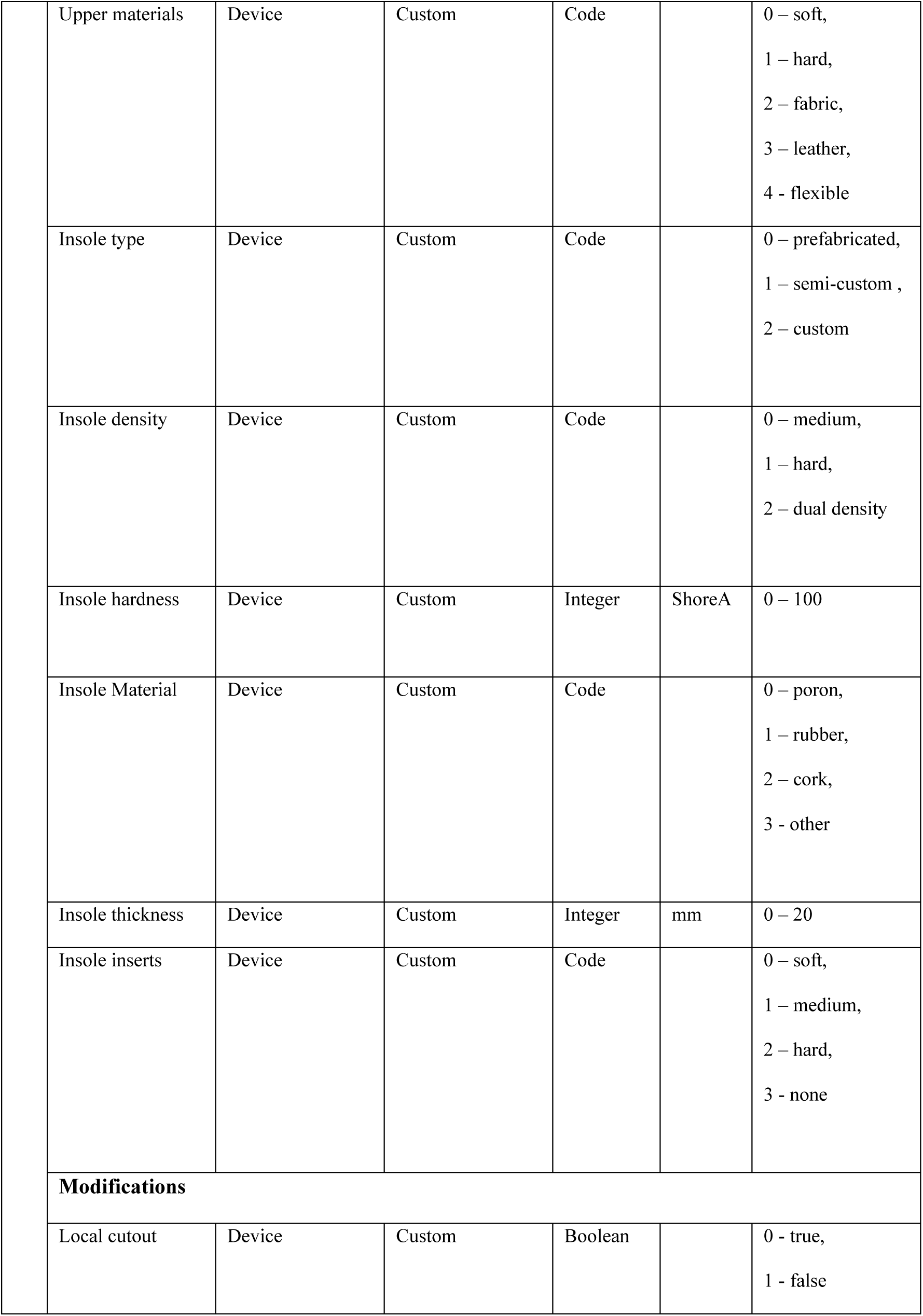
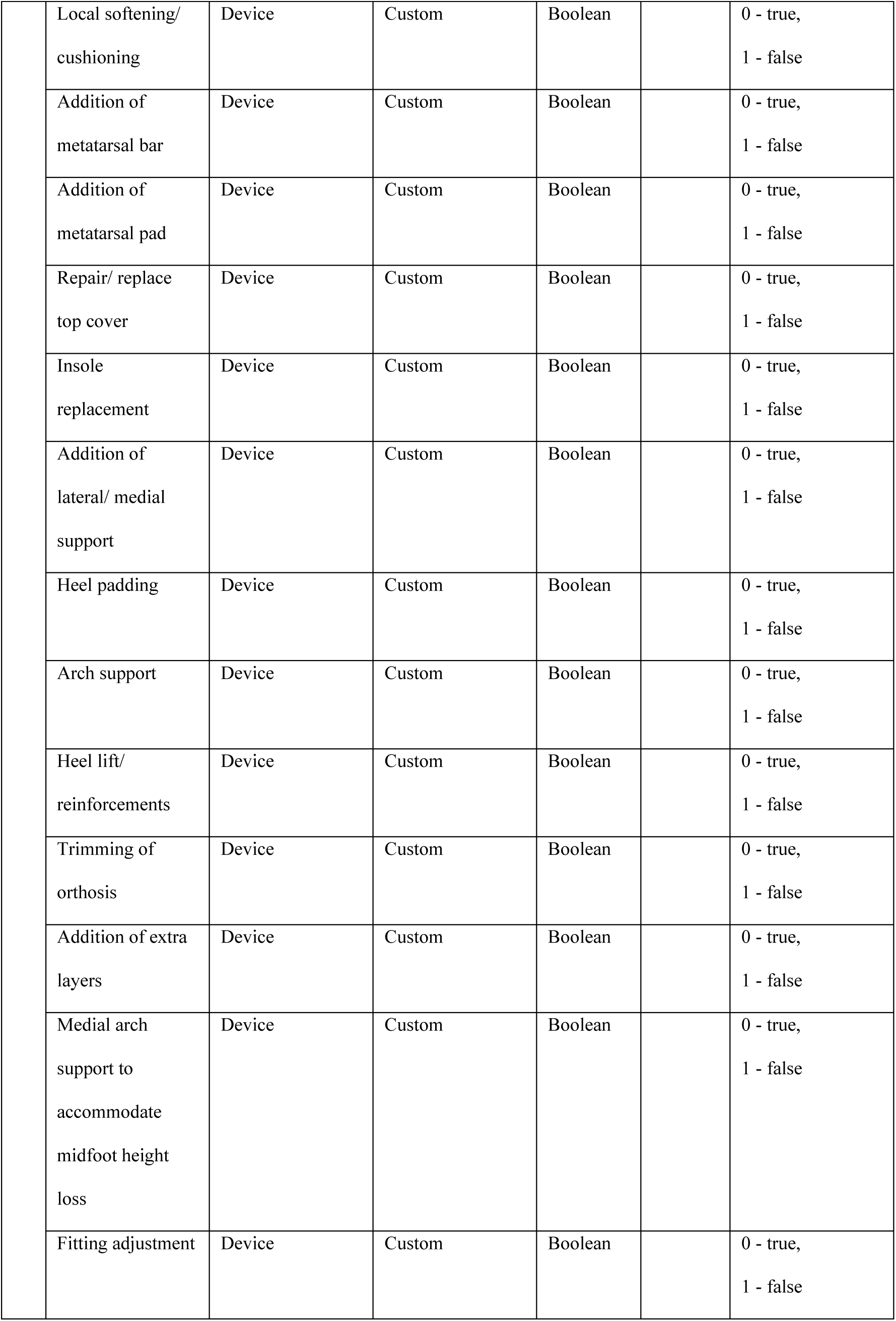
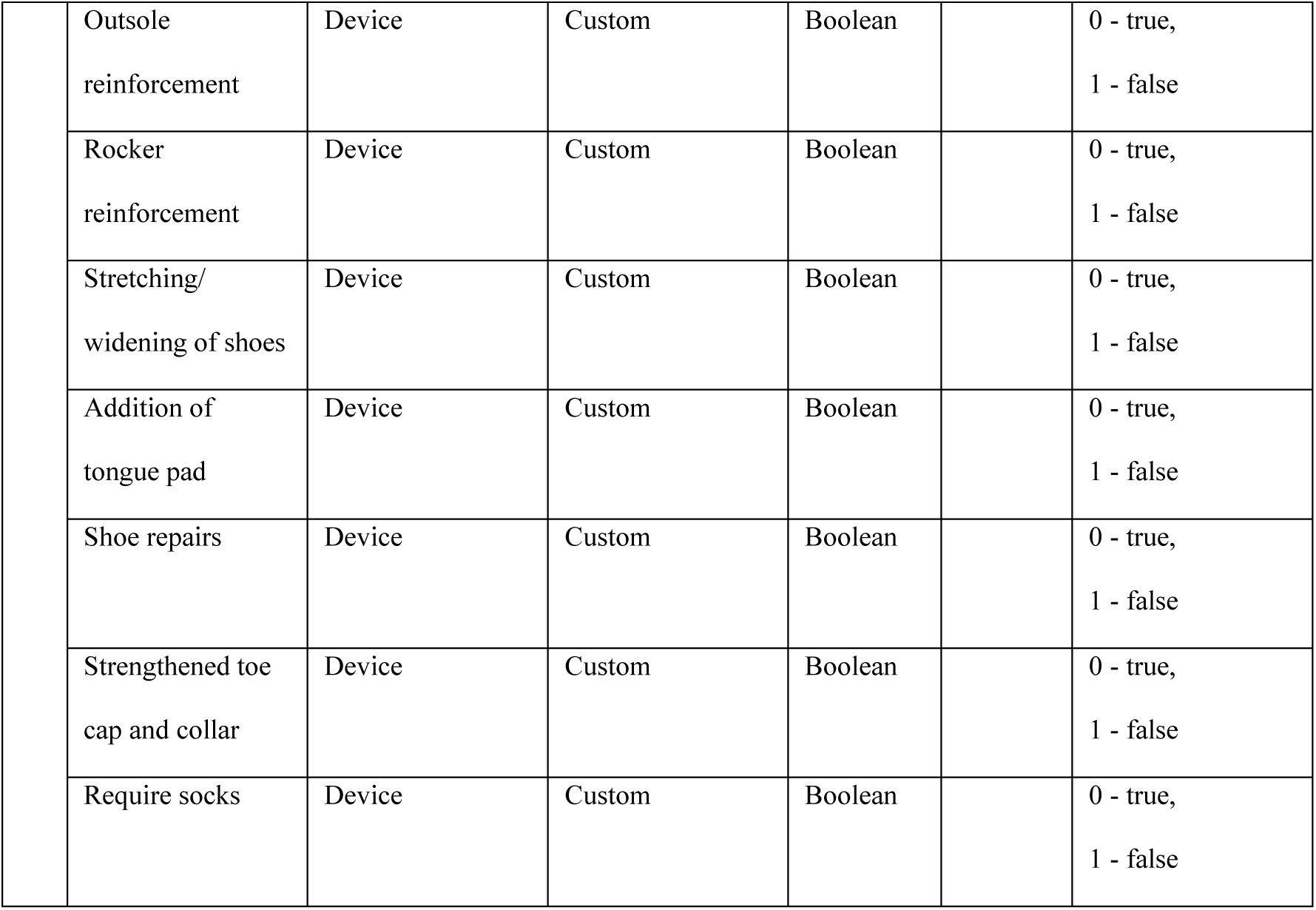
FHIR mappings of footwear prescription decision factors.

Therapy-related factors constitute the output group, encompassing footwear and insole specifications. These output variables are mostly discrete or Boolean, making them well-suited for multi-class classification and recommendation systems. Therapy related factors constitute the output group, encompassing footwear and insole specifications. These output variables are primarily discrete or Boolean, which makes them suitable for multi class classification and recommendation systems. Most modification variables are represented using Boolean values rather than detailed specifications or measurements. This approach reflects the framework’s focus on identifying the presence or absence of specific modifications to uncover patterns that support the decision-making process.

Including full configurations and exact measurements for each modification or footwear element would significantly increase the complexity of the dataset and create challenges for consistent data collection. For instance, footwear measurements are often captured using three-dimensional foot scanners. While these are important for the manufacturing process, the goal of this framework is to understand the type of footwear prescribed for specific patient conditions, rather than to capture precise measurements. Additionally, integrating devices like foot scanners into existing CDSSs may be impractical in many healthcare environments.

The integration of FHIR resources and standardized coding systems such as SNOMED CT and LOINC enhances interoperability. However, several variables rely on custom codes, reflecting the current lack of standardization in footwear prescription terminology—an area that presents opportunities for future development. Each variable’s datatype, unit, and sample values are clearly specified, offering practical guidance for data collection and preprocessing. It is important to note that while the sample values serve as a reference point, real-world data distributions may differ significantly.

In summary, Table 5 outlines a well-structured and comprehensive dataset schema that balances clinical detail with behavioral and contextual relevance. Further validation from clinical and industry experts is recommended to refine custom-coded fields and ensure applicability across diverse real-world scenarios.

## Discussion

### Principal Findings

This study presents a structured and interoperable framework for prescribing offloading footwear in the management of diabetic foot ulcers, designed to integrate into electronic health records and clinical decision support systems using FHIR standards. The schema captures a wide range of clinical, behavioural, environmental, and therapeutic factors, organized within a consistent structure informed by the WHO Dimensions of Adherence framework.

A central feature of the framework is its emphasis on semantic clarity. Redundant or overlapping factors are intentionally assigned to the most relevant domain by the WHO dimensions. For example, although footwear type may influence both clinical assessment and therapeutic planning, it is mapped solely within the therapy-related domain. This design choice improves clarity, prevents duplication, and ensures that the data can be applied consistently across longitudinal tracking systems and digital infrastructures.

In addition to enabling clinical documentation and decision support, the framework provides a foundation for AI and machine learning (ML) applications. Its structured categorization supports the development of high-quality datasets that can be used to develop CDSSs and train AI and ML models.

### Comparison With Prior Work

To the best of our knowledge, no previous study has systematically standardized or structured the decision factors involved in footwear prescription. Existing research has primarily focused on evaluating the effectiveness of offloading interventions, with limited attention to the underlying decision-making processes or their representation in digital systems. While clinical guidelines, such as those from the IWGDF on the Diabetic Foot, offer valuable principles, they lack the structure necessary for integration into electronic health records (EHRs) or computational tools such as CDSSs.

Recent work has begun to explore the application of artificial intelligence in this space. So far, only one study has conceptually proposed the idea of applying AI-powered clinical decision support system for personalized offloading device prescriptions, integrating patient-specific factors such as preferences, ulcer risk, and usage context to recommend tailored footwear and orthotics [4]. This study is among the first to conceptualize AI-driven personalization in DFU offloading footwear and presents an initial set of decision factors. However, the study focuses solely on forefoot ulceration and does not provide a structured approach to integrating in CDSSs or utilizing with AI and ML techniques.

The concept of AI and ML has been explored with smart and sensor-based footwear interventions. For example, a study presented the design and implementation of a real-time smart insole system that monitors plantar pressure and temperature using cost-effective, wearable sensors [106]. The study emphasizes home-based monitoring and the potential for integration with intelligent alert systems. While smart footwear has the potential to collect data and support the application of AI and ML models, its use in practice remains limited. This is largely due to the high cost of embedded sensors [107], which makes these interventions prohibitively expensive for many patients. This is one of the main reasons we excluded smart footwear interventions from the review, as their practical application remains limited.

Some studies have explored the ML techniques in orthotics. For example, a review highlighted the growing use of algorithms such as support vector machines, decision trees, and deep learning to optimize orthotic design and performance [108]. However, this review did not focus solely on orthotic interventions but also included prosthetic interventions. Another study conducted a case study that applied machine learning models to predict adherence to medical footwear based on behavioural and contextual variables, highlighting the importance of non-biomechanical factors [109]. Other empirical studies have applied ML model such as decision trees to prescribe orthotics for patients with Pes Planus [110] and deep learning models to prescribe orthotics that can improve patients’ gait [111].

While existing studies only discuss the feasibility of applying AI in footwear and orthotic contexts, they lack a standardized framework for organizing input and output factors. In contrast, this study offers a formal structure aligned with FHIR resources, designed to support both digital documentation and ML workflows. The therapy-related factors can serve as output for CDSSs or class labels for a dataset needed to train AL and ML modes. While patient-related, condition-related, socioeconomic, and health system-related can form the inputs of CDSSs or act as dependent variables in a dataset for AI and ML models. This dual-use model positions the schema for integration into real-world clinical decision support systems and intelligent predictive platforms.

### Limitations

The main objective of this scoping review was to identify and organize relevant decision factors, rather than to evaluate outcome effectiveness. As a result, the methodological quality of the included studies was not assessed. However, the relevance of the identified factors should be further evaluated through expert engagement, such as surveys and focus group discussions.

In addition, mapping these factors to FHIR resources should be guided by expert review and the use of standardized terminology systems. The proposed custom FHIR extensions will also require validation in real-world settings. It is important to note that some variables naturally span multiple domains, and rigid classification may not fully reflect their multidimensional relevance.

The proposed schema has not yet been tested in clinical workflows. Future work should include implementation studies to assess the usability of the framework, its integration into CDSSs, and its effectiveness in supporting real-time decision-making and predictive analytics.

## Conclusions

This scoping review systematically identified and organized clinical, behavioural, and contextual factors influencing the prescription of offloading footwear in the management of DFUs. By synthesizing evidence from a diverse body of literature, it developed a structured data schema aligned with the WHO Dimensions of Adherence and HL7 FHIR standards. This schema supports standardized data representation, facilitates integration into electronic health records, and provides a foundation for developing CDSSs and AI-driven tools.

While the review did not assess the methodological quality of the included studies, it prioritized breadth and comprehensiveness. Notably, it mapped 117 unique decision factors, 54% of which required custom FHIR extensions, offering a novel contribution to the field. The inclusion of socioeconomic and behavioural determinants underscores the importance of patient-centred approaches, particularly for enhancing adherence to prescribed footwear.

Future research should focus on validating the proposed schema through expert consensus, surveys, and focus group discussions. Implementation studies are also needed to evaluate its usability, effectiveness, and integration within CDSSs, with the goal of improving DFU management, reducing ulcer recurrence, and enabling personalized care at scale.

## Data Availability

All data is available in the manuscript.

## Author Contributions

KK conceptualized and designed the study, conducted the scoping review, performed data extraction and analysis, and led the manuscript drafting. MAK contributed to the study design, conceptual development, methodology, and visualization; supervised and administered the project; secured funding; and substantively revised the manuscript. LD contributed to the interpretation of findings and critically reviewed the manuscript with a focus on clinical implications. SA provided input on industry applications and offered insights into real-world implementation. MN participated in the review process and supported data extraction and analysis. All authors have read and approved the final manuscript and agree to be personally accountable for their own contributions and to ensure the accuracy and integrity of the work, even in areas where they were not directly involved.

## Ethics approval

NA

## Patient and public involvement

N/A

## Abbreviations

AI: Artificial intelligence
CDSS: Clinical decision support system
COP: Centre of pressure
DFU: Diabetes-related foot ulcer
DPN: Diabetic peripheral neuropathy
EHR: Electronic health records
FHIR: Fast Healthcare Interoperability Resources
HCP: Human centered design
HL7: Health Level Seven International
HRQoL: Health-related quality of life
IWGDF: International Working Group on the Diabetic
Foot ML: Machine Learning
PAD: Peripheral arterial disease
PGrad: Peak pressure gradient
PICO: Population intervention comparison outcome
PMap: Peak pressure map
PP: Plantar pressure
PPP: Peak plantar pressure
PTC: Peak pressure time curve
PTI: pressure time integral
PTM: Peak pressure time map
RCT: randomized controlled trial
SNOMED: Systematized nomenclature of medicine
LOINC: Logical Observation Identifiers Names and Codes
ICD-10: International Classification of Diseases, 10th Revision

